# Impaired contour object perception in psychosis

**DOI:** 10.1101/2024.07.02.24309795

**Authors:** Rohit S. Kamath, Kimberly B. Weldon, Hannah R. Moser, Samantha Montoya, Kamar S. Abdullahi, Philip C. Burton, Scott R. Sponheim, Cheryl A. Olman, Michael-Paul Schallmo

## Abstract

Contour integration, the process of joining spatially separated elements into a single unified line, has consistently been found to be impaired in schizophrenia. Recent work suggests that this deficit could be associated with psychotic symptomatology, rather than a specific diagnosis such as schizophrenia. Examining a transdiagnostic sample of participants with psychotic psychopathology, we obtained quantitative indices of contour perception in a psychophysical behavioral task. We found impaired contour discrimination performance among people with psychotic psychopathology (PwPP, n = 62) compared to healthy controls (n = 34) and biological relatives of PwPP (n = 44). Participants with schizophrenia (n = 31) showed impaired task performance compared to participants with bipolar disorder (n = 18). We also measured responses during an analogous task using ultra-high field (7T) functional MRI and found higher responses in the lateral occipital cortex of PwPP compared to controls. Using task-based functional connectivity analyses, we observed abnormal connectivity between visual brain areas during contour perception among PwPP. These connectivity differences only emerged when participants had to distinguish the contour object from background distractors, suggesting that a failure to suppress noise elements relative to contour elements may underlie impaired contour processing in PwPP. Our results are consistent with impaired contour integration in psychotic psychopathology, and especially schizophrenia, that is related to cognitive dysfunction, and may be linked to impaired functional connectivity across visual regions.

## 1. Introduction

Atypical visual perception in schizophrenia, and in psychotic psychopathology more broadly, are well documented (Butler et al., 2008; Phillips & Silverstein, 2013). Anomalies include abnormal contrast sensitivity (Calderone et al., 2013), surround suppression (Schallmo et al., 2015), integration of visual contours (Silverstein et al., 2009), processing of motion (Chen, 2011), and facial emotion recognition (Turetsky et al., 2007). Disruptions in visual functioning in schizophrenia are associated with greater symptom severity, including higher rates of psychotic symptoms such as delusions and hallucinations (Keane et al., 2018), and increased cognitive disorganization (Phillips & Silverstein, 2003). Examining basic perceptual functions that are well understood in the healthy visual system may provide insight into the etiology of visual disturbances in schizophrenia and other disorders of psychotic psychopathology. This motivated us to examine visual contour perception across groups representing a spectrum of severity of psychotic symptoms.

Contour integration is a visual function that is important for object and scene perception. This process involves the linking of spatially separated elements into unified edges or shapes, and plays an important role in detecting boundaries and subsequently separating objects from their backgrounds. Contour integration follows the Gestalt laws of proximity, continuity, and similarity, such that contour elements which conform to these principles are more easily integrated (Loffler, 2008). Contour processing occurs as early as primary visual cortex (V1), in which the responses of individual orientation-tuned neurons to contour elements within their classical receptive fields are modulated by nearby elements outside the receptive field (e.g., collinear facilitation; Angelucci & Bressloff, 2006). This modulation depends on a combination of feed-forward (e.g., thalamocortical), lateral (e.g., horizontal connections within V1), and feed-back mechanisms (Angelucci et al., 2017), and is thought to involve reciprocal processing between lower regions that process basic visual features (e.g., V1), and higher object or category selective visual areas such as the lateral occipital complex (LOC; Fang et al., 2008). This notion is supported by functional MRI (fMRI) studies in human observers, showing that both V1 and higher visual areas such as LOC play important functional roles in contour processing (Altmann et al., 2003; Murray et al., 2002; Qiu et al., 2016).

Several studies have reported abnormal contour object processing in PwPP. Many of these have presented contours of various shapes within a field of noise Gabor elements, and asked participants either to report the location of the contour (e.g., quadrant or side of the screen; Uhlhaas et al., 2005), or to report the configuration of a contour shape (e.g., pointing left or right; Silverstein et al., 2012). Contour discrimination thresholds can be measured by adding orientation jitter to the contour elements to perturb the alignment of each target Gabor element relative to the axis of the overall shape. Participants with schizophrenia have been shown to have less tolerance to jitter among contour elements (Pokorny, Lano, et al., 2021), and this deficit may be representative of PwPP more broadly (Grove et al., 2018). Further, there is some evidence to suggest that even individuals with schizotypal traits (but without a diagnosed psychotic disorder) show less tolerance to jitter in contour integration tasks (Panton et al., 2018). Less tolerance for jitter has been reported in people with psychotic disorders for both closed (Keane et al., 2016; Silverstein et al., 2012) and open contours (Robol et al., 2013; Schallmo et al., 2013). Others have reported decreased contour discrimination accuracy in psychosis, but no difference in jitter thresholds (Moran et al., 2022; Silverstein et al., 2015). Neuroimaging studies of contour and shape integration in schizophrenia have suggested abnormalities at various levels of the visual processing hierarchy (Keane et al., 2021; Pokorny, Espensen-Sturges, et al., 2021), including regions such as LOC (Silverstein et al., 2009, 2015). Perceptual closure deficits in schizophrenia may also be linked to abnormal functional connectivity within and between visual areas (van de Ven et al., 2017), including LOC (Li et al., 2020).

Psychotic illnesses such as schizophrenia, schizoaffective disorder, and bipolar I disorder have a substantial genetic component to their etiology (Uher & Zwicker, 2017), with evidence suggesting that schizophrenia has very high heritability (e.g., 79%; Hilker et al., 2018). The study of endophenotypes, quantifiable neuro-behavioral traits that can be shared between people with psychotic illnesses and their first-degree biological relatives (Iacono, 2018), allows for an examination of the potential role of genetic liability in abnormalities associated with psychosis. Although relatively few studies have examined the role of genetic liability for psychosis in contour perception, there is some evidence to suggest that contour perception deficits may be more closely linked with disease processes than to genetic liability for psychotic illnesses (Pokorny, Lano, et al., 2021; Schallmo et al., 2013).

To examine abnormal visual contour perception across a spectrum of psychotic psychopathology, including individuals with genetic liability for psychosis, we undertook the current study as part of the Psychosis Human Connectome Project (P-HCP). We acquired data from 3 groups: PwPP, healthy control participants, and a sample of first-degree biological relatives; these are individuals who share roughly 50% of their genes with someone having a diagnosed psychotic illness. Full methodological details on the larger study are available in our previous publications (Demro et al., 2021; Schallmo et al., 2023). We used both psychophysical behavioral measures and ultra-high field (7T) fMRI to examine contour object processing in psychosis. For the latter, we defined visual regions of interest (ROIs) representing the contour stimuli and evaluated whether their responses and connectivity between the ROIs depended on the perceptibility of contours and the presence of psychotic psychopathology. Using both psychophysical and fMRI measures allowed us to map anomalies in contour perception onto psychotic phenomenology, and assess their neural correlates across types of psychotic psychopathology (i.e., schizophrenia, schizoaffective disorder, bipolar disorder with psychosis, first-degree biological relatives of PwPP). Our 7T fMRI methods allowed us to measure these neural anomalies with the highest degree of anatomical precision achieved to date. Our results suggest that reciprocal processing between low and intermediate / higher level visual areas (i.e., between V1 and LOC) may play an important role in abnormal contour processing in psychosis.

## 2. Method

### 2.1 Participants

A total of 140 adult participants between the ages of 18 and 60 were recruited to participate in our psychophysical and 7T fMRI experiments as a part of the Psychosis-Human Connectome Project. Five of the participants did not complete the 7T fMRI portion of the study (i.e., behavioral psychophysics data only), leaving 135 participants with functional neuroimaging datasets. Of the 140 total participants, 62 were people with a psychosis spectrum disorder, 44 were first-degree biological relatives without psychotic psychopathology, and 34 were healthy control participants with no family history of psychosis (see *Table 1* for demographic information). Recruitment and demographic information from the same study have also been reported in recent publications from our group (Demro et al., 2021; Ramsay et al., 2020; Schallmo et al., 2023; *Table 1*). A subset of PwPP (n = 37) were recruited to return for a second ‘repeat’ scanning session, in which they repeated the same psychophysical and 7T fMRI experiments.

**Table 1:**
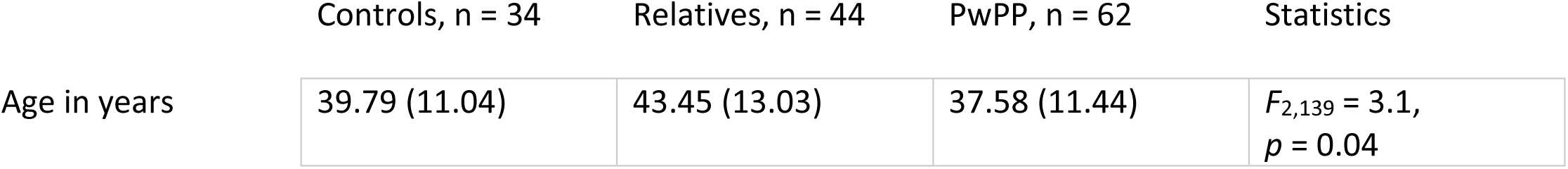
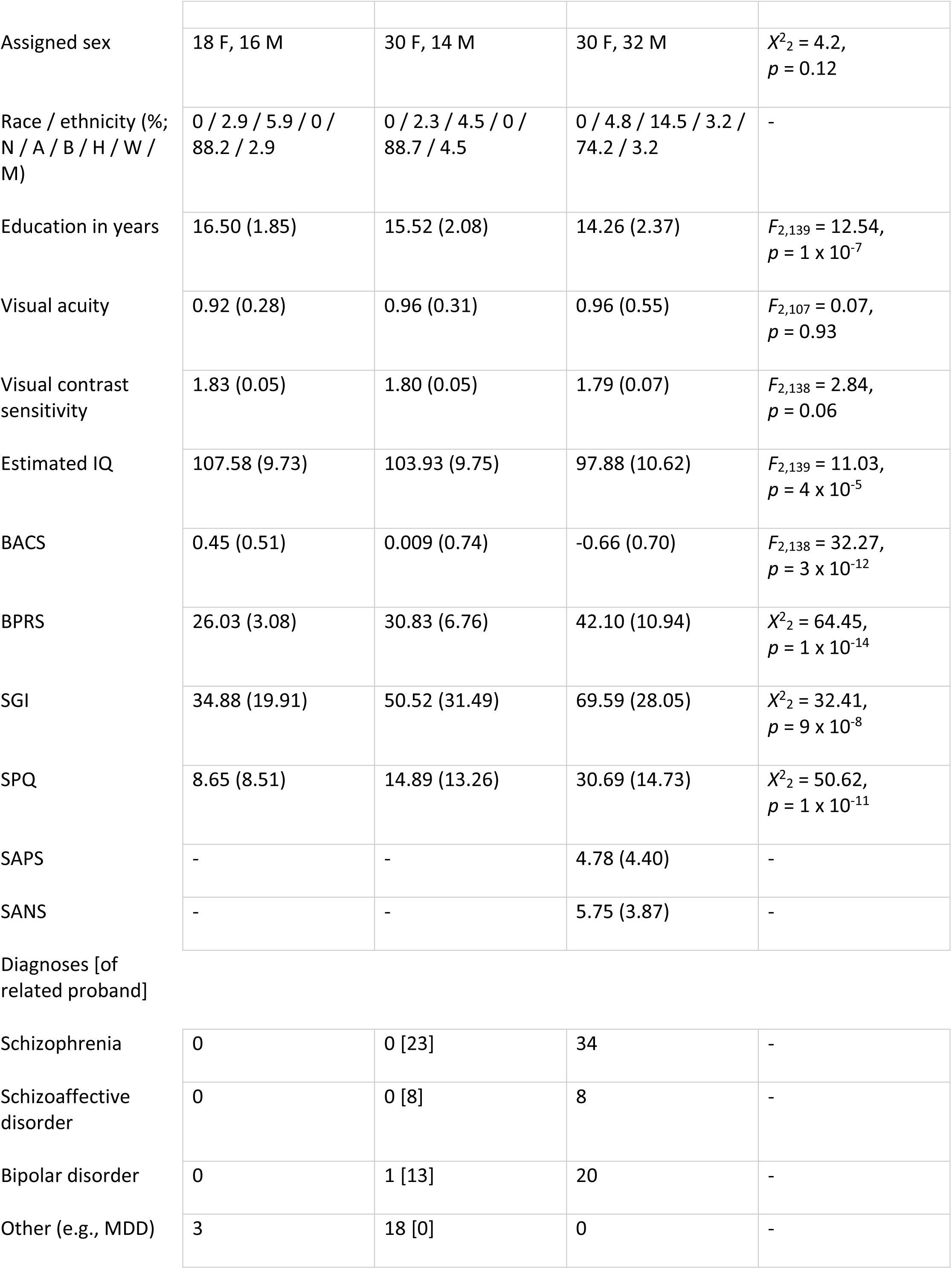
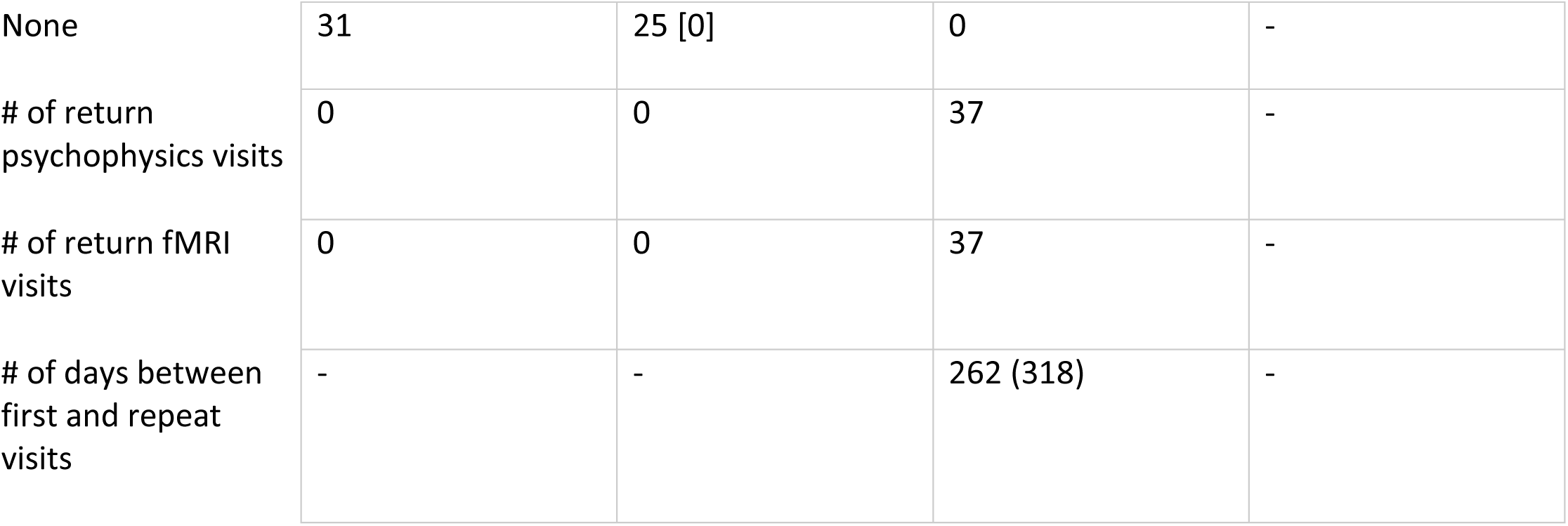
Participant demographics, cognitive, and symptom measures. Data are presented as mean (standard deviation), unless otherwise noted. Racial and ethnic designations, as defined by the National Institute of Health, are abbreviated as follows: A = Asian or Pacific Islander, B = Black (not of Hispanic origin), H = Hispanic, N = Native American or Alaskan Native, W = White (not of Hispanic origin), M = More than 1 race or ethnicity, or other. Visual acuity was assessed with a Snellen eye chart (Snellen, 1862); the decimal fraction is reported (e.g., 0.5 indicates 20/40). Estimated IQ was assessed using the Wechsler Adult Intelligence Scale (WAIS-IV) (Wechsler, 2012). BACS = Brief Assessment of Cognition in Schizophrenia, Z score (Keefe et al., 2004), BPRS = Brief Psychiatric Rating Scale (Ventura et al., 2000), SGI = Sensory Gating Inventory (Hetrick et al., 2012), SPQ = Schizotypal Personality Questionnaire (Raine, 1991), SANS = Scale for the Assessment of Negative Symptoms (Andreasen, 1989), SAPS = Scale for the Assessment of Positive Symptoms (Andreasen, 1990). Diagnoses were based on the Structured Clinical Interview for DSM-IV-TR disorders (SCID; M. First et al., 2002). For the relative group, the number of individuals related to probands with a particular psychotic disorder diagnosis is listed in square brackets.

PwPP were stable, outpatient individuals with a confirmed clinical diagnosis of a psychotic illness. Research diagnoses for all participants were made by trained clinicians with expertise in psychosis spectrum disorders using the Structured Clinical Interview for DSM-IV-TR (American Psychiatric Association, 2000; M. B. First & Gibbon, 2004). Consensus diagnoses for PwPP were made by at least 2 doctorate-level clinical psychologists with expertise in psychosis spectrum disorders.

Participants were screened for having normal or corrected-to-normal vision and no neurological conditions or learning impairments. Participants were excluded for having any current or past nervous system disease, for experiencing loss of consciousness greater than 30 minutes, or if they had any condition that made it difficult for them to lie still within an MRI scanner or complete any study tasks. Participants were excluded on the day of their visit if they reported having consumed more than two alcoholic beverages or any illicit or recreational drugs during the previous 24 hours, or if they had any conditions or implants that would prevent safe MR scanning (e.g., claustrophobia, incontinence, or specific implanted devices). All participants had completed two 3T MRI scanning sessions (approximately 2.5 hours total) prior to 7T scanning, and had fewer than 20% of volumes from 3T fMRI excluded by a motion censoring algorithm (threshold = 0.5 mm of framewise displacement).

Participants were recruited from the Minneapolis Veterans Affairs Medical Center, the Fairview Riverside Hospital in Minneapolis, and at outreach programs organized by the University of Minnesota Department of Psychiatry. They provided written informed consent prior to participation and were either compensated $150 per complete visit or $20 per hour for incomplete visits. Participant ability to provide informed consent was determined using the University of California Brief Assessment of Capacity for Consent (UBACC; Jeste et al., 2007). All procedures were approved by the Institutional Review Board at the University of Minnesota (IRB #1607M0981) and conformed to the guidelines for research on human subjects from the Declaration of Helsinki.

### 2.2 Clinical Measures

Clinical and cognitive assessments were carried out by trained staff members under the supervision of a post-doctoral associate and trained clinicians with expertise in the care and evaluation of individuals with psychosis spectrum disorders. A clinical battery comprising several items was administered to all participants. Items included the Brief Psychiatric Rating Scale (BPRS; Overall & Gorham, 1962; Ventura et al., 2000), with subscale items taken from Wilson & Sponheim (Wilson & Sponheim, 2014), the Sensory Gating Inventory (SGI; Hetrick et al., 2012), the Wechsler Adult Intelligence Scale-IV (WAIS-4; Wechsler, 2012), and the Brief Assessment of Cognition in Schizophrenia (BACS; Keefe et al., 2004). Visual acuity was also measured using the Snellen Acuity Measurement Chart (Snellen, 1862). PwPP additionally completed the Scales for Assessment of Negative Symptoms and Positive Symptoms (SANS & SAPS; Andreasen, 1989, 1990). The BPRS, SANS, and SAPS were acquired with a focus on clinical functioning over the past 30 days. These measures were reacquired at each 7T research visit, if more than 30 days had elapsed since the participant’s previous visit. Other measures were acquired at a separate clinical visit.

### 2.3 Visual Display and Stimuli

Psychophysical experiments were carried out on an Apple Mac Pro with an Eizo FlexScan SX2462W monitor (refresh rate = 60 Hz) using PsychoPy (version 1.85.2; Peirce, 2007). Monitor mean luminance was 60.8 cd/m^2^. A chin rest was used to stabilize head position, and viewing distance was set at 70 cm.

Visual stimuli (*Figure 1*) were designed to mirror those that have been previously used by Silverstein and colleagues (Silverstein et al., 2012). These stimuli comprised of a grid of 170 Gabor elements 14° visual angle wide by 11.3° tall. Gabor elements had a spatial frequency of 5 cycles per degree and a Gaussian envelope with *SD* = 0.067°. There were 2 cycles present in each ∼0.4° wide (6 *SD*) Gabor, although only the central 1 cycle had high contrast. Fifteen Gabor elements near the center of the display formed an egg-shaped contour object (5.9° wide, 4.68° high) that either pointed towards the left or the right. Gabors within the contour object had a minimum spacing of 1.09° and a maximum spacing of 1.13°. The size of the contour shape was set at 5.9° wide by 4.7° tall. One thousand exemplar stimulus grids were procedurally generated and saved for the presentation in the task. To manipulate the detectability of the contour object stimulus, we jittered the relative orientation of each Gabor element with respect to the axis of the egg (see below). One hundred and fifty-five Gabor elements composed the background, with a minimum spacing of 0.8° visual angle. Signal-to-noise, defined as the average spacing between adjacent background elements divided by the average spacing between adjacent contour object elements was 0.87. Pilot data from an early version of the task with different stimulus parameters were not included in the current study (for full details, see Schallmo et al., 2023).

**Figure 1:**
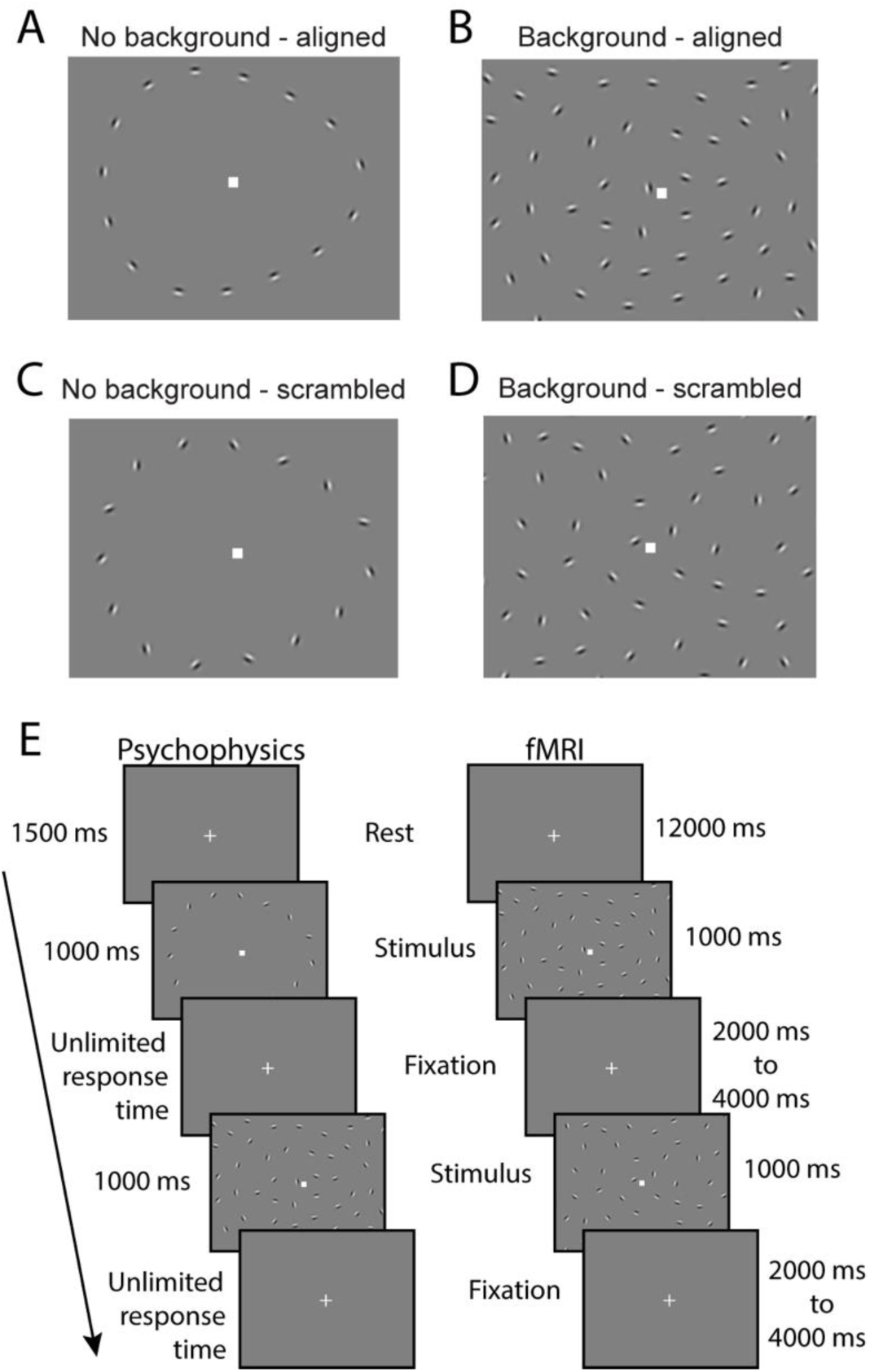
Task paradigm and example stimuli used for each of the 4 conditions. **A-D)** Examples of contour stimuli used for each of four conditions. **E)** Trial sequences are shown for the psychophysics experiment and the fMRI experiment. Egg-shaped contours composed of 15 Gabor elements were presented with or without background elements. Participants reported which direction the egg pointed while maintaining central fixation.

FMRI experiments used the same task stimuli, which were presented within the scanner. Stimuli were presented on a screen at the back of the scanner bore using an EPSON projector with a 60 Hz refresh rate and a mean luminance of 271 cd/m^2^, and viewed through a mirror mounted on the head coil. Viewing distance was 100 cm.

### 2.4 Experimental Procedure

#### 2.4.1 Behavioral Psychophysics

Participants performed a behavioral psychophysics experiment before their fMRI scan, on the same day. Task stimuli consisted of three types of trials, with the lowest difficulty consisting of a scrambled (45° jitter) contour object without any background Gabor elements (*Figure 1C*). This was used to ensure participants could accurately perceive the directionality of the egg without a background, even when the contour elements did not perfectly line up. The intermediate difficulty stimuli consisted of aligned (0° jitter) contour objects within a field of background elements (*Figure 1B*). This allowed us to measure the accuracy with which participants could discriminate the directionality of a perfectly aligned egg with background Gabor elements. The greatest difficulty consisted of jittered contour objects within a field of background elements. The alignment of the contour elements was manipulated to quantify the highest degree of orientation variability, or jitter threshold, at which the subject could accurately discriminate the directionality of the egg contour. Jitter was manipulated using a Psi adaptive staircase method implemented within PsychoPy. Jitter was varied in increments of 3° (range 0-45°) to identify the particular jitter angle at which each participant could perceive the direction of the egg with 70% accuracy (i.e., jitter threshold).

In the psychophysical task, participants were instructed to fixate on a square at the center of the screen, and use their peripheral vision to discern whether the egg-shaped contour object was pointing towards the left or the right. Trials began with this central fixation square for 1500 ms, followed by the presentation of the stimulus for 1000 ms, and a response period during which participants were asked to use the arrow keys on the keyboard to indicate their answer (*Figure 1E*). Response time was not limited. Two blocks were administered, with each block consisting of 3 interleaved staircases of 30 trials each. Each block also included 20 trials of the scrambled contour stimuli with no background and 20 trials of the aligned contour stimuli with a background (260 trials total across the two blocks). The order of trials was randomized within each block. Participants were allowed to take a self-timed rest in between the two blocks, and were also told that they could take a short pause during a block if needed by withholding their response until they were ready to proceed. Each block lasted about 5 minutes, and total task duration was approximately 10 minutes.

#### 2.4.2 Functional MRI

Our fMRI paradigm was designed to measure brain responses to the contour object stimulus. This fMRI version of the contour object task was analogous to our psychophysical task, with the following differences. The fMRI paradigm consisted of six main experimental blocks (each 24 s). Each block contained six stimulus presentation events (trials; 1000 ms duration). In four of these trials per block, aligned contour stimuli (*Figure 1A & B*) were presented. For the other two trials in each block, the contour stimuli were completely scrambled (i.e., fixed at 45° jitter; *Figure 1C & D*). During 3 of the 6 blocks, contours were presented with background Gabor stimuli (*Figure 1B & D*), while in the other three blocks, contours were presented without background elements (*Figure 1A & C*). The degree of orientation jitter in the aligned with background condition was controlled by a 3-up, 1-down adaptive staircase (García-Pérez, 2000). Jitter level began at 0° (completely aligned) and increased in increments of 3°. Jitter in the aligned without background fMRI trials matched that in the preceding aligned with background block. The inter-stimulus interval was randomized between 2-4 s. Between trials, the fixation mark (white square) was presented on a mean gray background (i.e., background Gabors were not present). The block order alternated between background and no background, beginning with background. There were an additional 12 sec of rest (i.e., no stimuli) before and after each of the 6 main experiment blocks. A single fMRI task run lasted 5.2 or 7.8 min in total (see below), and each subject completed 2 fMRI runs within a single scanning session.

In the fMRI task, participants were asked to fixate on a central fixation square and report the direction in which the egg-shaped contour pointed. If they were unsure what direction the egg pointed (e.g., in the scrambled contour with background trials, in which the egg was not easily perceived), they were told to make their best guess. Participants were asked to respond as soon as possible after the stimulus presentation using a 4 button MR-compatible response device (Current Designs, Philadelphia, PA).

The fMRI paradigm also included a functional localizer condition, which consisted of repeated presentations of a dynamic egg-shaped contour stimulus, with contrast reversing at a frequency of 2 Hz (*Figure 2A*). This condition was designed to identify the retinotopic regions in early visual cortex that represented the spatial position of the egg stimuli, and consisted of alternating blocks of rest and stimulation. A 12 s block of rest was presented first, to allow the fMRI signal to reach a steady state. Data from this first rest block were discarded during analysis. This was followed by a 12 s block of stimulation during which the directionality of the egg contour changed randomly between left and right every 2 s (*Figure 2A*). In total, 7 blocks of rest and 6 blocks of stimulation were administered in alternating order. The functional localizer condition was 2.6 min in total duration, and was presented immediately before the first main experiment block as part of the first of the two fMRI runs (7.8 min total duration for the first fMRI run).

**Figure 2:**
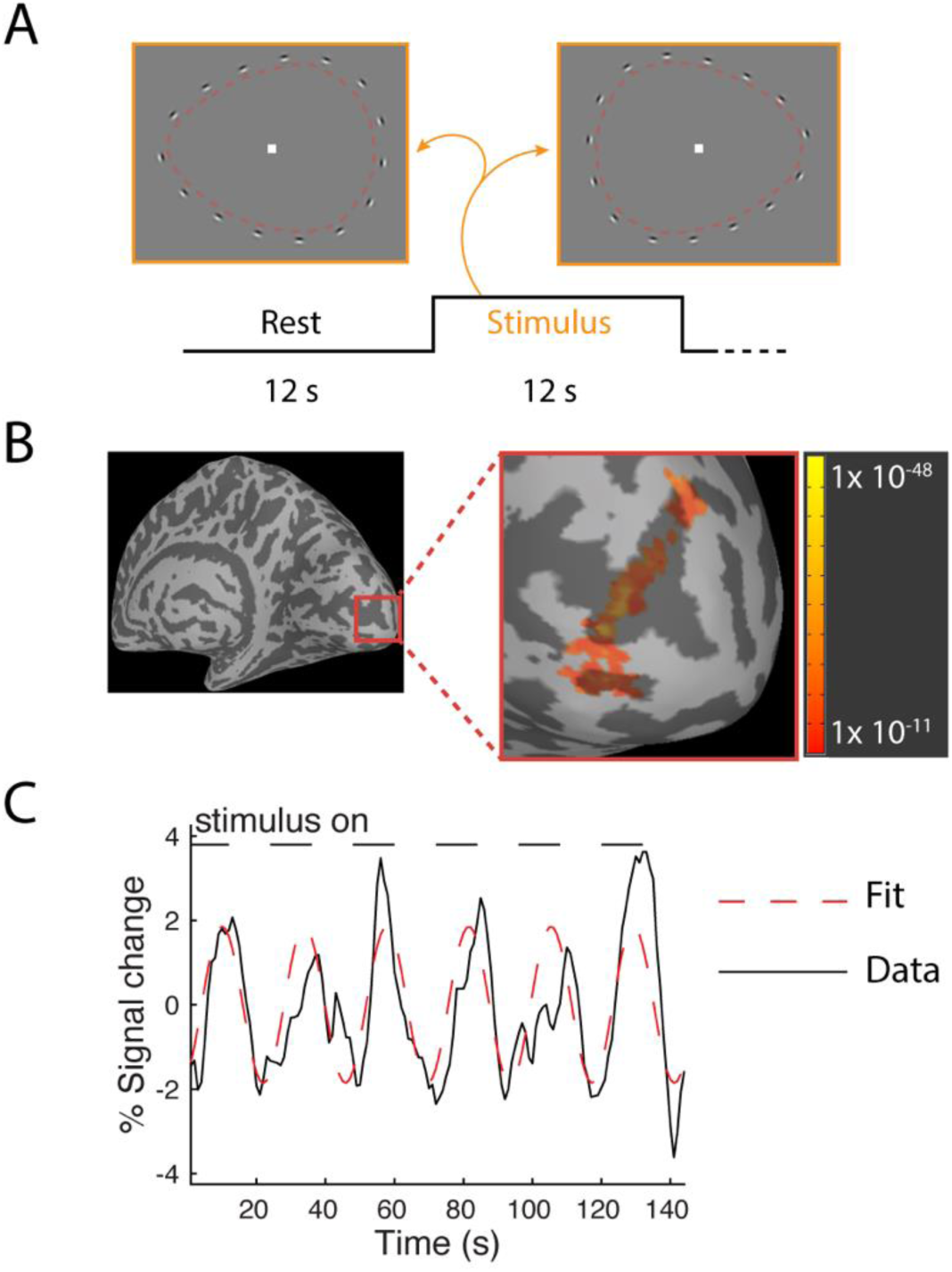
FMRI functional localizer. **A)** Example of block design used to define retinotopic region within V1 that represented the contour object (red dashed lines for visualization only). **B)** Representative V1 ROI drawn utilizing functional localizer data. Color bar shows *p* value for associated ROI. **C)** Fourier analysis of the functional localizer data; the best fitting sine wave at the stimulus presentation frequency is overlaid on a representative plot of the fMRI response.

MRI data were acquired on a Siemens MAGNETOM 7 Tesla scanner (software version updated from VB17 to VE12U in July, 2019). Gradient echo fMRI data were collected using a Nova Medical radiofrequency head coil (1 transmit and 32 receive channels) with 1.6 mm isotropic resolution in 85 slices (1 s repetition time [TR], 22.2 ms echo time [TE], 45° flip angle, anterior-posterior phase encode direction, parallel imaging acceleration factor = 2, multi-band acceleration factor = 5). There were 468 TRs during the first contour object perception task fMRI run, and 312 TRs in the second run. A single run (3 TRs) was acquired at the beginning of the scanning protocol with an opposite phase encode direction (posterior-anterior) to facilitate correction for distortion due to B_0_ inhomogeneity (Schallmo et al., 2021). Additionally, we placed 5 mm thick dielectric pads (3:1 calcium titanate powder in water, by mass) under the neck and beside the temples, as this has been shown to improve B_1_ transmit homogeneity in the cerebellum and temporal lobe regions during 7T MRI (Vu et al., 2015).

### 2.5 Data Processing and Analysis

#### 2.5.1 Psychophysics data processing

Psychophysical data were analyzed in MATLAB (version 2016a; MATHWORKS, Natick, MA) using the Palamedes toolbox (Prins & Kingdom, 2018). Data from two conditions were used as catch trials; jittered contour stimuli without any background elements, and aligned contour stimuli with background elements. Participants who did not achieve 85% accuracy on the former trial type or 72.5% accuracy on the latter were excluded from further analyses of threshold data, and only catch trial data were analyzed from these participants. This was done to ensure that threshold data were only examined for participants from whom we could expect to obtain meaningful threshold estimates, i.e., they understood and could perform the task with no background elements, and could achieve sufficiently high accuracy (≥ 72.5%) for aligned contours with background stimuli to justify quantifying jitter thresholds. Threshold data from 3 participants (3 PwPP) were excluded for low accuracy in the no background catch trials, whereas threshold data from another 13 participants (1 control, 5 first-degree relatives, 7 PwPP) were excluded for low accuracy in the with-background catch trial condition. These exclusion criteria were defined post-hoc based on an examination of the distributions across all participants. A contingency table analysis was carried out to determine whether data sets from one participant group (e.g., PwPP) were being excluded more often than would be expected by chance, compared to other groups. We saw no significant difference in exclusion rates across groups [*X^2^* _(2, *n =* 138)_ = 2.09, *p* = 0.35].

Jittered contour discrimination data across the three separate staircases within each block were pooled together for fitting purposes, yielding two independent jitter threshold estimates (one for each block) per data set. Contour discrimination thresholds were calculated by fitting the staircase data from the jittered contour condition with background stimuli using a Weibull function (*Supplemental Figure 1*). Guess rate was fixed at 50%. Lapse rates for each session in each participant were estimated using the accuracy from the aligned contour condition (i.e., 0° jitter), with a minimum lapse rate of 2.5%. Aligned contour data were also included with the jittered contour staircase data for fitting purposes. Neither our choice to estimate individual lapse rates, nor to include aligned contour data during fitting had a qualitative effect on our pattern of results (data not shown). However, these steps did tend to yield subjectively better psychometric function fits overall. Thresholds were calculated as the jitter angle at which participants performed with 70% accuracy, based on the fit psychometric function. Threshold estimates that were between -3° and 0° (i.e., one jitter step) were set to 0°, as these were judged to be within an acceptable margin of measurement error. A total of 6 threshold estimates were adjusted in this way. Estimated threshold values below -3° or above 90° (i.e., outside the valid range) were excluded. A total of 17 threshold estimates were excluded in this way. These exclusion criteria were established *a priori*.

#### 2.5.2 fMRI data processing

At the individual subject level, functional MRI data were processed primarily in AFNI (version 18.2.04; (Cox, 1996). This included gradient nonlinearity correction (using *gradunwarp*, version 1.0.3; <u>github.com/Washington-University/gradunwarp</u>), motion correction, distortion compensation, and alignment to an anatomical T_1_ weighted scan from a 3T scanner acquired in a separate session. The T_1_ weighted anatomical data were processed in FreeSurfer (version 5.3; Fischl, 2012) as part of the HCP Minimal Preprocessing Pipeline (version 3.22.0; Glasser et al., 2013). Following the above corrections, fMRI data were smoothed with a 2 mm full-width, half-maximum Gaussian kernel and masked to exclude non-brain regions, then scaled to percent signal change. A generalized linear model (GLM) analysis was used to estimate response magnitudes (beta weights) for each of the four stimulus conditions using AFNI’s *3dDeconvolve* function. Data for the four main experiment conditions were analyzed in an event-related fashion, using the gamma function provided within AFNI as the assumed shape of the hemodynamic response function. Motion censoring was carried out by removal of all TRs where head motion exceeded 0.5 mm. Further, the six motion parameters estimated during motion correction were included as nuisance regressors in the design matrix, as well as three Legendre polynomials for detrending slow fluctuations in the fMRI signal.

FMRI data from individual participants were retroactively excluded if they exceeded a limit of 0.5 mm of head motion on greater than 20% of TRs during the two 7T fMRI scanning runs; 1 control and 1 PwPP were excluded in this way. FMRI data sets were also excluded if the participant failed to make any response (correct or incorrect) on more than 10% of the trials during the fMRI task runs; 1 control, 5 relatives, and 5 PwPP were excluded due to poor fMRI task performance. These exclusion thresholds were defined *a priori*.

Regions of interest (ROIs) were defined in each hemisphere at the individual subject level. ROIs were defined in primary visual cortex within retinotopic regions that represented the position of the egg-shaped contour object using the functional localizer data. Significantly activated regions were identified using a Fourier analysis of the fMRI time series data during the functional localizer condition by examining the response at the stimulus presentation frequency (5 cycles / scan; *Figure 2B & C*). ROIs were defined by identifying significantly activated voxels with a threshold coherence value of 0.3 (*F*_2,142_ = 30.4, uncorrected *p* = 1 x 10^-11^) or higher. Coherence is similar to an unsigned correlation value, and is a standard metric for Fourier time series analyses of fMRI data (Engel et al., 1997; Schallmo et al., 2016). We further restricted our ROIs to voxels within V1 with phase values in the range of -3.14 to -1.96 and +2.75 and +3.14; phase values are circular between -π to π, and reflect the offset between the observed fMRI time course during the functional localizer condition and the stimulus presentation timing, due to the sluggish hemodynamic response (*Figure 2C*). Cluster-wise *p*-value correction (Cox et al., 2017; Eklund et al., 2016) was performed using AFNI’s *3dClustSim*, with all ROIs satisfying a minimum cluster-corrected threshold of *p* < 0.01. ROIs were defined within V1 using an anatomically defined V1 mask in FreeSurfer (Wang et al., 2015). The V1 mask was dilated by two steps and eroded by one step using AFNI’s *3dmask_tool* to fill in any holes within the volumetric surface.

Lateral occipital complex (LOC) ROIs were defined using the same Fourier analysis results which intersected with an anatomically defined LOC ROI (areas LO1 and LO2 combined) from the FreeSurfer atlas (Wang et al., 2015). ROI positions were then verified through visualization on an inflated FreeSurfer white matter surface using AFNI’s surface mapping program, SUMA (Saad et al., 2004). Lateral geniculate nucleus (LGN) ROIs were defined by a 2 mm sphere positioned based on anatomical landmarks (i.e., gray matter region lateral to the medial geniculate nucleus and cerebral peduncles, ventral to the pulvinar) using the structural T_1_ data. Definition of each ROI (V1, LOC, and LGN) for each individual participant was not always successful, resulting in different numbers of datasets for each ROI (*Supplemental Table 1*). For each participant, average fMRI response amplitudes were extracted from each ROI for each of the 4 main experimental conditions based on the results of the GLM analysis above for further analyses within MATLAB.

#### 2.5.3 Beta series analysis

We performed a beta series analysis (Cisler et al., 2014) to quantify task-based functional connectivity between our ROIs. Beta weights were estimated from every voxel separately for each individual trial using the *stim_timesIM* flag in AFNI’s *3dDeconvolve* command in an otherwise identical GLM analysis to the one described above. The voxel-wise average of these first-level beta weight time series was taken from a seed ROI, and a second-level GLM analysis was performed, using the extracted seed beta weights from each trial as predictors (i.e., in the design matrix). Second-level beta weights were then extracted from a target ROI and averaged across voxels. These second-level beta weights provided a directional (i.e., seed to target), but not causational, measure of task-based functional connectivity between our seed and target regions.

### 2.6 Statistics

Group-level statistical analyses were performed in MATLAB. *F*-test statistics were obtained using repeated measures analyses of variance (ANOVAs), with participants treated as a random effect. Data were compared across groups; for our fMRI results, we also included stimulus background and contour alignment as factors in our analyses. Normality and homogeneity of variance were assessed by visual inspection of the data. In cases where these assumptions were not met, non-parametric Kruskal-Wallis (K-W) tests were used. Where appropriate, two sample *t*-tests were used to examine post-hoc differences between groups and/or conditions. Correlational values (*r*) presented throughout are Pearson’s correlation coefficients (two-tailed, unless noted), except for correlations with visual acuity data, for which Spearman ranked correlation was used to account for the skew in the data. A threshold of *p* = 0.05 was used throughout to determine statistical significance. We used two-way random effects intraclass correlation coefficients (also called ICC (3, k); Koo & Li, 2016) to quantify variability within and across experimental sessions.

### 2.7 Data availability

Data for this study are available through the National Data Archive (nda.nih.gov/edit_collection.html?id = 3162).

## 3. Results

Using a contour object perception task, we characterized behavioral performance as well as 7T fMRI responses within different visual brain areas in a group of 62 individuals with psychosis, 44 first-degree biological relatives, and 34 healthy controls. In addition to GLM analyses of the fMRI response, we performed beta series analyses to quantify task-based functional connectivity between visual areas.

### 3.1 Behavioral data

#### 3.1.1 Contour discrimination accuracy

Participants completed a contour discrimination task in which they reported whether an egg-shaped contour composed of Gabor elements pointed to the left or right. Contour discrimination accuracy was assessed in two stimulus conditions: scrambled contours with no background, and aligned contours with background stimuli. Across all participants, contour discrimination accuracy was high for scrambled contours without background elements (stimuli shown in *Figure 1C*; median accuracy = 100%; *Figure 3A*), and for aligned contours with background stimuli (shown in *Figure 1B*; median accuracy = 92.50%; *Figure 3B*). Near-ceiling performance was expected for these conditions, and shows that in general participants understood the task instructions and could reliably discriminate contour direction. Participants who did not reach criterion level accuracy in either of these two conditions (horizontal dashed lines in *Figure 3A & B*) were excluded from further analyses (see Methods for full details).

**Figure 3:**
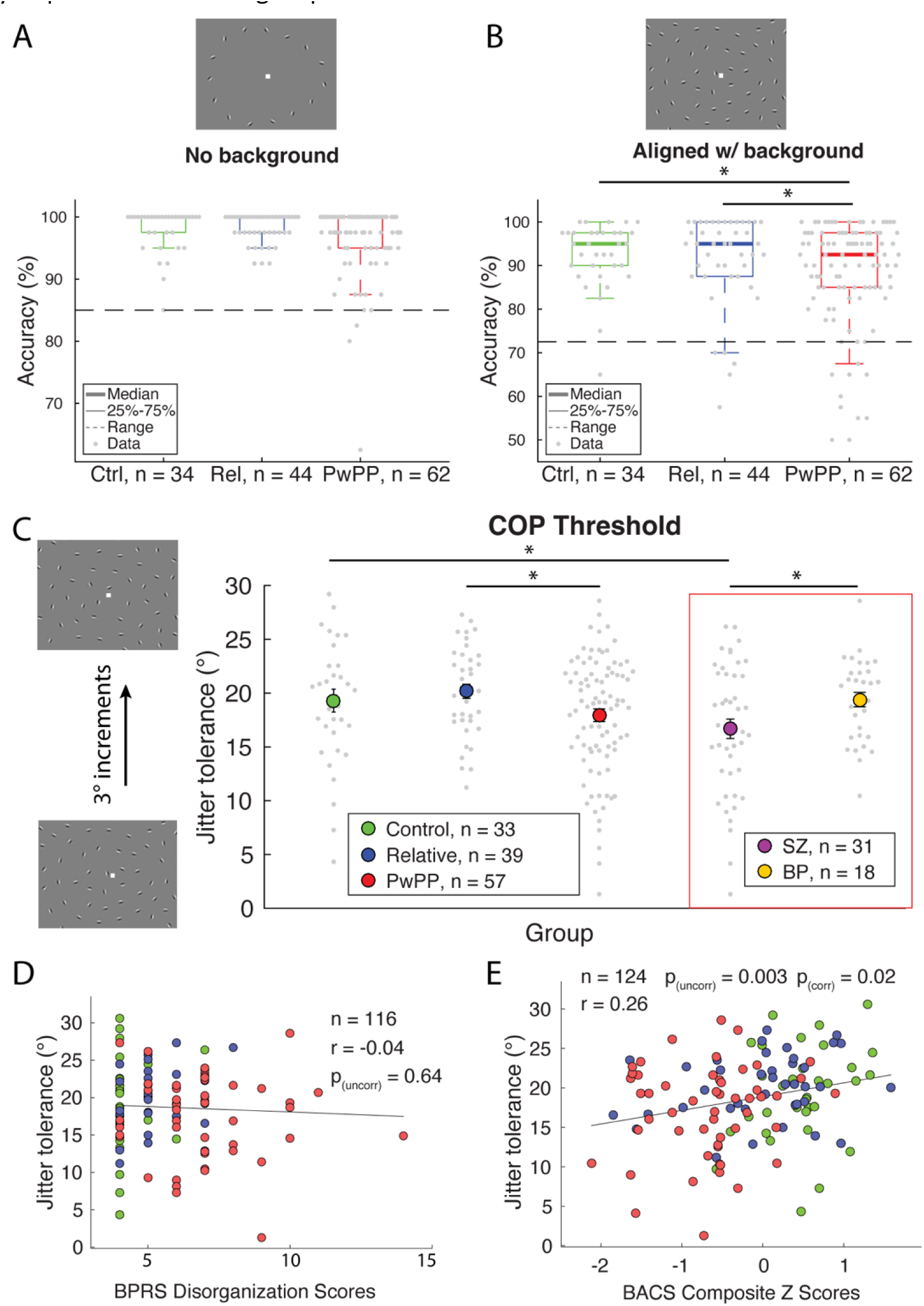
Behavioral task results. Contour discrimination accuracy for **A)** scrambled contour stimuli without background elements, and **B)** aligned contour stimuli with background elements. Thick line shows the group median, boxes show interquartile range, whiskers show 1.5 x interquartile range, gray dots show all data points. **C)** Jitter tolerance across groups. Data are presented as individual data points (gray) alongside group average (color). Data points within red box are from subsets of the PwPP group. Error bars show ± 1 *SEM*. Relationship between jitter thresholds and **D)** score on the BPRS disorganization subscale or **E)** BACS *Z* score. Pearson’s correlation (*r*) values are shown as uncorrected (uncorr), and Bonferroni corrected values (corr). Asterisks indicate significant differences across participant groups (*p* < 0.05).

We next compared contour discrimination accuracy across our three subject groups. There were no significant differences in accuracy across the 3 groups for the scrambled contour stimuli without background elements (i.e., the lowest-difficulty condition; *Figure 3A*; K-W test, main effect of group; *Χ*^2^_2_ = 0.93, *p* = 0.6). However, there was a significant difference across groups for aligned contour stimuli with background elements (intermediate difficulty; *Figure 3B*; K-W test, main effect of group; *Χ*^2^_2_ = 10.7, *p* = 0.004), with PwPP showing significantly lower accuracy than healthy controls (K-W test, main effect of group; *Χ*^2^ = 7.21, *p* = 0.007), and first-degree relatives (*Χ*^2^ = 7.58, *p* = 0.005). This result provides support for the notion that contour detection accuracy is impaired among individuals with psychosis.

As relatively few studies have examined contour perception across various psychotic disorders, we also explored whether contour discrimination accuracy differed across individuals with different psychosis spectrum diagnoses (i.e., schizophrenia and bipolar disorder). No significant differences in accuracy were observed between these psychosis diagnostic groups and healthy controls for the scrambled contour stimuli without background elements (main effect of group; *Χ*^2^_2_ = 0.62, *p* = 0.7; *Supplemental Figure 2*). However, there was a significant difference across these groups for aligned contour stimuli with background elements (main effect of group; *Χ*^2^ = 11.29, *p* = 0.004; *Supplemental Figure 2*), driven by lower accuracy among participants with schizophrenia vs. controls (*Χ*^2^ = 11.68, *p* = 6 x 10^-4^), suggesting that contour discrimination accuracy may be particularly impaired within this group.

#### 3.1.2 Jitter thresholds

Contour discrimination thresholds were measured by fitting adaptive staircase data from the jittered contour with background condition using a psychometric function. Thresholds reflect the jitter angle at which participants discriminated the direction of the egg-shaped contour (left or right) with 70% accuracy. Across all participants, jitter thresholds were generally high (mean = 18.90°) but varied substantially across individuals (*SD* = 5.76°). We observed a significant difference in jitter thresholds between groups (ANOVA, main effect of group; *F* _(2, 126)_ = 3.6, *p* = 0.02). PwPP showed similar tolerance for jitter compared to healthy controls (*F* _(1, 88)_ = 2.32, *p* = 0.13), but significantly less tolerance compared to first-degree relatives (*F* _(1, 94)_ = 6.97, *p* = 0.009; *Figure 3C*). Thresholds differed significantly across psychosis spectrum diagnoses (ANOVA, main effect of group; *F* _(2, 79)_ = 3.62, *p* = 0.031); participants with schizophrenia showed significantly lower jitter thresholds compared to healthy controls (ANOVA, main effect of group; *F* _(1, 62)_ = 4.69, *p* = 0.034) and participants with bipolar disorder (ANOVA, main effect of group; *F* _(1, 47)_ = 5.55, *p* = 0.023; *Figure 3C*). Lower tolerance for orientation jitter during contour discrimination among PwPP, and especially individuals with schizophrenia, is consistent with impaired contour integration in these groups (Grove et al., 2018; Keane et al., 2012, 2016; Moran et al., 2022; Pokorny, Lano, et al., 2021; Robol et al., 2013; Schallmo et al., 2013; Silverstein et al., 2012, 2015).

We next examined the two independent jitter threshold estimates from the two experimental blocks in each participant, and found that test-retest reliability within the psychophysical session was high (ICC = 0.81; *Supplemental Figure 4A*). A subset of our PwPP (N = 37 out of 66, 56%) also returned for a repeat session several months after their initial experimental session (see *Table 1*). Thus, we were able to additionally examine jitter threshold estimates across time within this subset of our PwPP, and found that there was substantial variability in threshold estimates within the same individuals over a period of several months (ICC = 0.39; *Supplemental Figure 4B*). Together, this indicates that jitter threshold estimates were reliable within an experimental session, but that thresholds within individuals from our psychosis group varied across time, suggesting they may reflect the current state of perceptual and cognitive functioning, rather than a stable individual trait.

#### 3.1.3 Relation to clinical measures

A primary goal for our study was to examine how contour discrimination may be related to the clinical manifestations of psychotic illness. To this end, we examined correlations between jitter thresholds and scores for disorganized symptoms (from the BPRS; Wilson & Sponheim, 2014), as well as positive symptoms (from SAPS), abnormal sensory experiences (from SGI), schizotypy (from SPQ), cognitive performance (BACS *Z* scores), and visual acuity. These measures were chosen *a priori*; disorganization was examined because previous findings have suggested a relationship between the level of disorganization symptoms and contour perception (Silverstein & Keane, 2011), while SAPS, SPQ and SGI were chosen due to our primary interest in psychotic symptomatology. BACS scores allowed us to examine well known cognitive deficits in PwPP that may be associated with contour perception (Grove et al., 2018). Likewise visual acuity has been shown to differ among PwPP (Hayes et al., 2019) and may play an important role in contour perception (Keane et al., 2015). No significant correlations between jitter thresholds and BPRS disorganization, SAPS, or visual acuity measures were observed in our participants (*Figure 3*, *Supplemental Figure 3*; |*r*|-values < 0.08, uncorrected *p*-values > 0.5). This suggests that these measures associated with psychosis may not be closely related to performance in our contour discrimination task. However, we did observe a significant positive correlation between BACS *Z* scores and jitter thresholds (*Figure 3*; *r* _(122)_ = 0.26, uncorrected *p* = 0.003; Bonferroni corrected *p* = 0.02), indicating that poorer cognitive performance may be associated with less tolerance for orientation jitter during contour discrimination. Additionally, we observed significant correlations between lower jitter thresholds and higher SPQ scores as well as higher SGI scores, which did not survive corrections for multiple comparisons (*Supplemental Figure 3*; |*r*|-values = 0.19, uncorrected *p*-values < 0.04; Bonferroni corrected *p*-values > 0.05).

### 3.2 Imaging data

#### 3.2.1 Primary visual cortex

We first examined 7T fMRI responses within the retinotopic regions of primary visual cortex (V1) that represent the spatial position of the contour (*Figure 4A*) based on fMRI responses to the independent functional localizer (see Methods). We saw a main effect of contour in the V1 fMRI response, with scrambled contours producing slightly higher fMRI responses compared to aligned stimuli (ANOVA, main effect of contour; *F* _(1, 106)_ = 9.15, *p* = 0.003). V1 fMRI responses for contour stimuli presented within a field of background elements were higher than for contours presented without background elements (ANOVA, main effect of background; *F* _(1, 106)_ = 517.71, *p* < 0.001; *Figure ^4^B*); this was expected given the increased number of visual elements. We saw no significant overall differences in V1 fMRI responses across groups (ANOVA, main effect of group; *F* _(2,106)_ = 0.6, *p* = 0.6), and there were no significant interactions between group, contour alignment, or background (*F*-values < 0.53, *p*-values > 0.2; *Figure 4C*), indicating similar patterns of univariate V1 fMRI responses across our 3 participant groups. We also saw no significant group differences when comparing participants with schizophrenia, participants with bipolar disorder, and healthy controls (*F*-values < 1.42, *p*-values > 0.2; *Supplemental Figure 5*).

**Figure 4:**
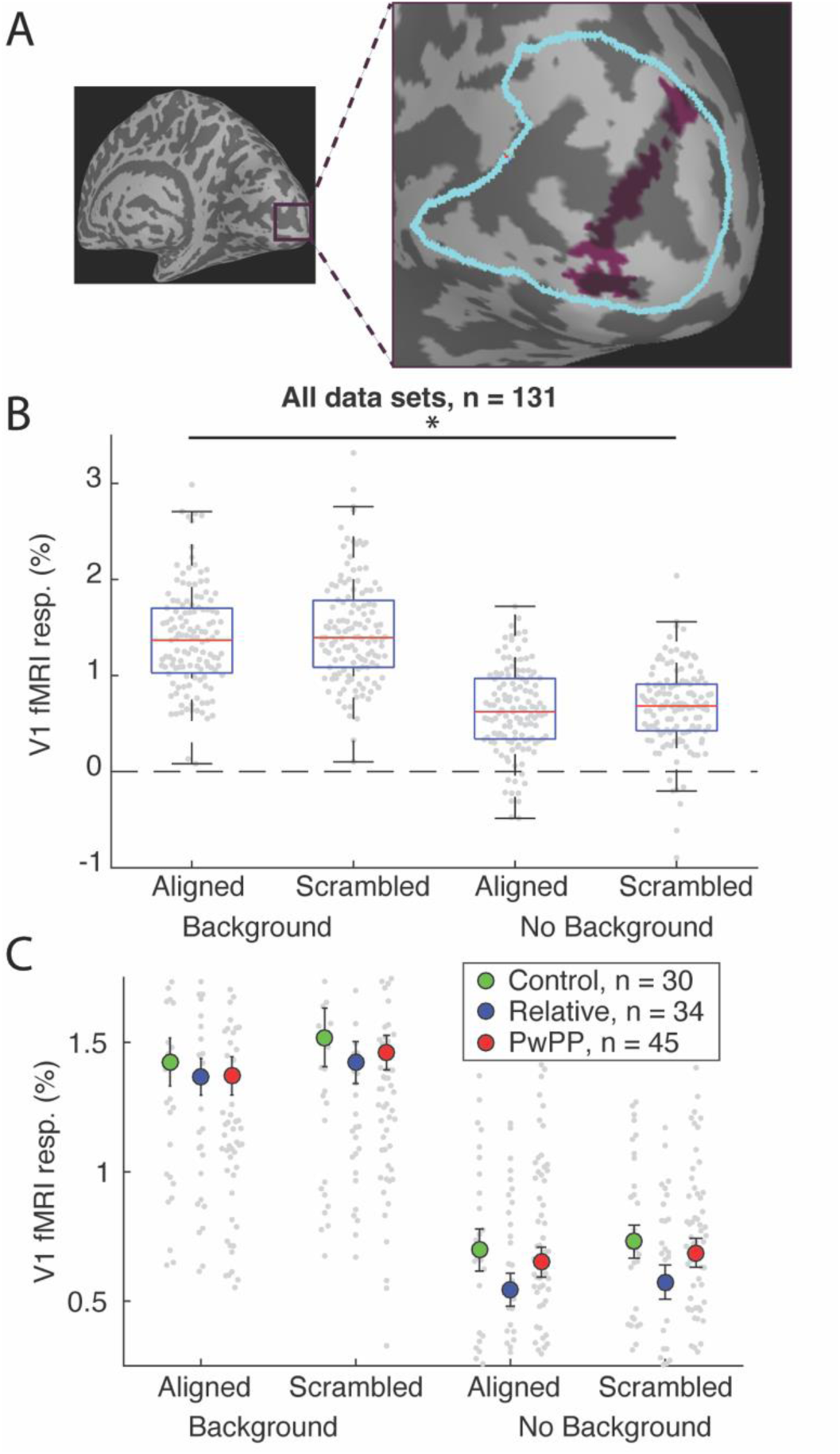
V1 fMRI results. **A)** Representative V1 contour ROI. Outline of anatomical V1 (Wang, 2015) shown in cyan. **B)** V1 fMRI responses across 4 stimulus conditions. Red line shows the median, boxes show interquartile range, whiskers show 1.5 x interquartile range, gray dots show all data points. **C)** V1 fMRI responses across participant groups. Small gray dots show individual subject data, larger circles show group means. Not all individual data points are shown in C, to better visualize the group means. Error bars show ± 1 *SEM*. Asterisk indicates significant difference across background vs. no background conditions (*p* < 0.05).

#### 3.2.2 Lateral geniculate nucleus

Next, we examined 7T fMRI responses from the lateral geniculate nucleus of the thalamus (LGN), which is the primary target for axons projecting from the retinae through the optic nerve, and in turn provides feed-forward input to the primary visual cortex. Unlike for V1, LGN ROIs were defined anatomically, and were not restricted based on retinotopic responses to the functional localizer (*Figure 5A*). LGN fMRI responses did not vary significantly with contour alignment (i.e., aligned vs. scrambled; ANOVA, main effect of contour; *F* _(2, 107)_ = 0.12, *p* = 1.0). However, fMRI responses in the LGN were significantly larger for contours presented with versus without background stimuli (ANOVA, main effect of background; *F* _(1, 107)_ = 82.97 *p* < 0.001; *Figure ^5^B*) as expected, given the increased number of visual elements. First-degree relatives showed overall higher fMRI responses in the LGN across stimulus conditions, as compared to healthy controls and PwPP (ANOVA, main effect of group; *F* _(2, 107)_ = 7.97, *p* = 6 x 10^-4^). There were no significant interactions between group, contour alignment, or background (*F*-values < 1.07, *p*-values > 0.3; *Figure 5*C). Group differences for LGN responses between controls, participants with schizophrenia, and participants with bipolar disorder were not significant (*F*-values < 1.79, *p*-values > 0.17; *Supplemental Figure 5*).

**Figure 5:**
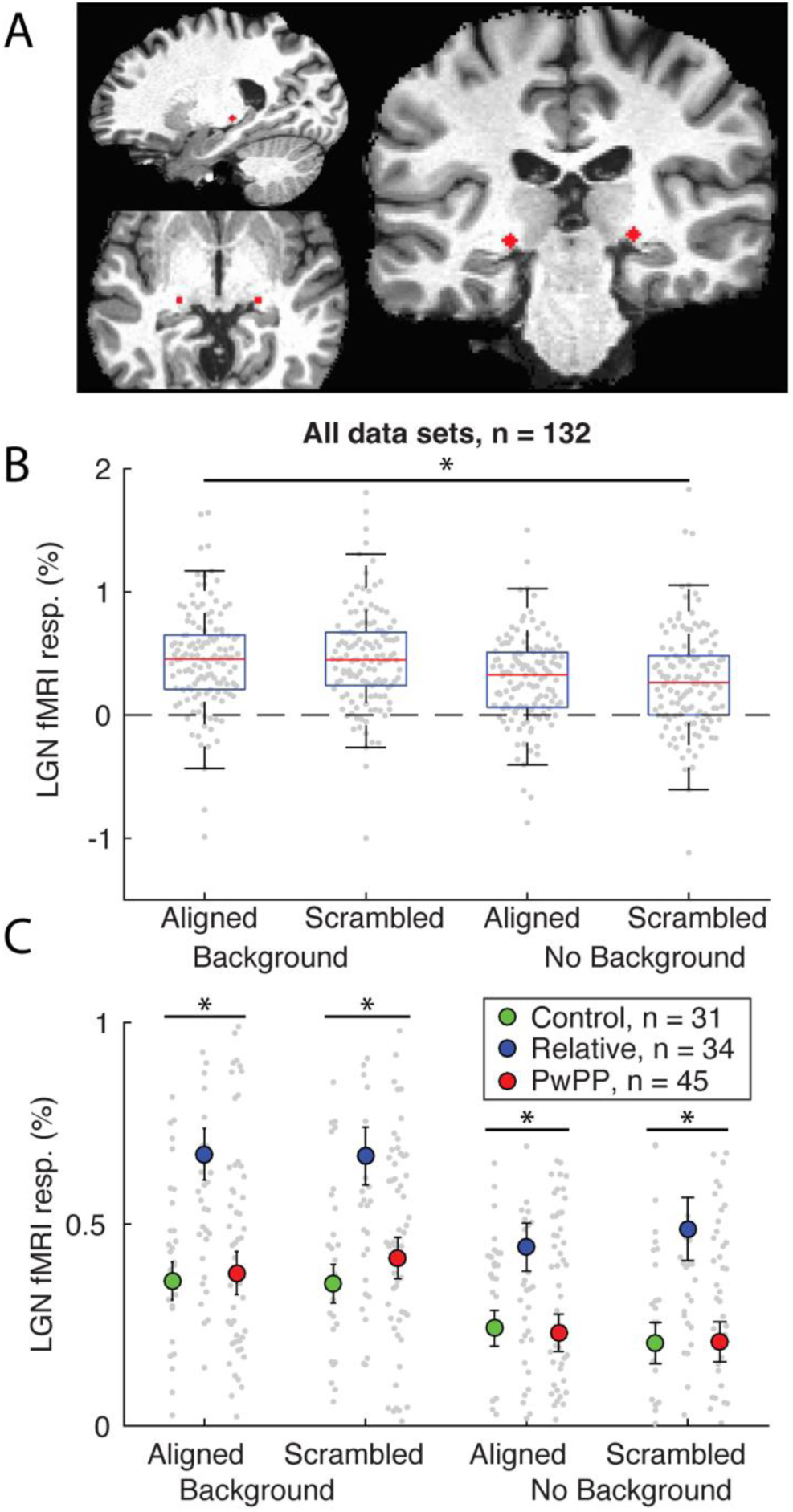
LGN fMRI results. **A)** Representative LGN ROI. **B)** LGN fMRI responses across 4 stimulus conditions. Red line shows the median, boxes show interquartile range, whiskers show 1.5 x interquartile range, gray dots show all data points. **C)** LGN fMRI responses across participant groups. Small gray dots show individual subject data, larger circles show group means. Not all individual data points are shown in C, to better visualize the group means. Error bars show ± 1 *SEM*. Asterisks indicate significant differences across background vs. no background conditions in B, and participant groups in C (*p* < 0.05).

#### 3.2.3 Lateral occipital complex

We also examined 7T fMRI responses within the lateral occipital complex (LOC), a region involved in the representation and perception of visual objects (Grill-Spector et al., 2001). LOC ROIs were defined based on fMRI responses in the lateral occipital cortex to the functional localizer (see Methods; *Figure 6*A). We saw no significant overall difference in the LOC fMRI responses for aligned versus scrambled contours (ANOVA, main effect of contour; *F* _(1, 104)_ = 2.96, *p* = 0.08). As in V1 and the LGN, fMRI responses in LOC were significantly higher for contours presented within a field of background elements versus contours presented alone (ANOVA, main effect of background; *F* _(1, 104)_ = 100.95, *p* < 0.001; *Figure 6B*). There was a significant contour by background interaction, such that fMRI responses were larger for aligned versus scrambled contour stimuli with background elements, but without background elements, responses were smaller for aligned versus scrambled contours (ANOVA, contour by background interaction; *F* _(1, 104)_ = 11.89, *p* = 8 x 10^-4^; *Figure 6B*). This result is similar to previous findings from fMRI studies of contour processing in areas V1 and V2 (Qiu et al., 2016), and appears consistent with the notion that the LOC is involved in detecting shapes within noisy stimulus displays (i.e., figure-ground segmentation). We saw no significant overall differences in LOC fMRI responses between groups (ANOVA, main effect of group; *F* _(2, 104)_ = 0.62, *p* = 0.5). However, PwPP showed higher fMRI responses versus controls when background elements were present, with relatives showing an intermediate response (ANOVA, group by background interaction; *F* _(2, 104)_ = 5.5, *p* = 0.005; *Figure 6C*). Similar results were obtained when comparing LOC responses across participants with schizophrenia, participants with bipolar disorder, and healthy controls (group by background interaction; *F* _(2, 66)_ = 4.3, *p* = 0.017; *^S^upplemental Figure ^5^*). However, differences between participants with schizophrenia vs. participants with bipolar disorder were not significant (*F*-values < 3.7, *p*-values > 0.06). Higher LOC fMRI responses to contours with backgrounds (but no difference without background stimuli) may suggest abnormal figure-ground processing among PwPP and relatives, such as a failure to adequately suppress the response to background elements.

**Figure 6:**
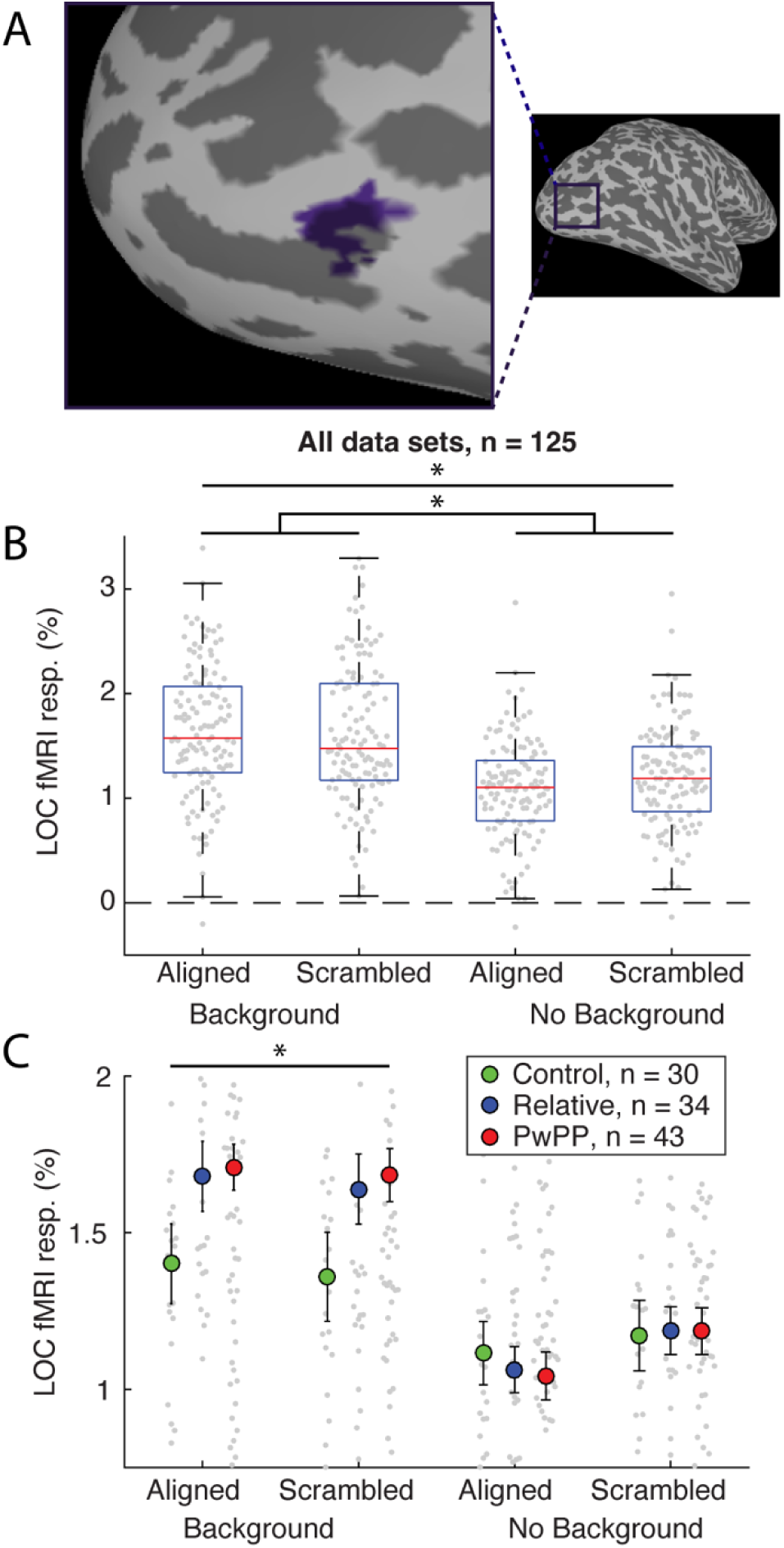
LOC fMRI results. **A)** Representative LOC ROI. **B)** LOC fMRI responses across 4 stimulus conditions. Red line shows the median, boxes show interquartile range, whiskers show 1.5 x interquartile range, gray dots show all data points. **C)** LOC fMRI responses across participant groups. Small gray dots show individual subject data, larger circles show group means. Not all individual data points are shown in C, to better visualize the group means. Error bars show ± 1 *SEM*. Asterisks indicate significant differences across background vs. no background conditions and a contour by background interaction in B, and a group by background interaction in C (*p* < 0.05).

#### 3.2.4 Task-based functional connectivity between LGN and V1

Previous work has suggested that people with psychosis (especially those with schizophrenia) may have aberrant functional connectivity between the thalamus and cortex (Woodward et al., 2012), and that such dysconnectivity may be important for understanding the etiology of psychotic symptoms. To examine thalamocortical functional connectivity during our contour object perception task, we performed a task-based functional connectivity analysis (beta series method; Cisler et al., 2014; see Methods) of our 7T fMRI data from V1 and the LGN. Although the second-level beta weights from this analysis provide a directional measure of functional connectivity (i.e., from a seed region to a target region), we cannot infer anything about *causality* in this relationship (i.e., we cannot say that signals from the seed region *cause* changes in the response within the target ROI). For this reason, we performed our analyses in both directions, with the LGN and V1 each serving as the seed in one analysis and the target in the other analysis (*Figure 7*).

**Figure 7:**
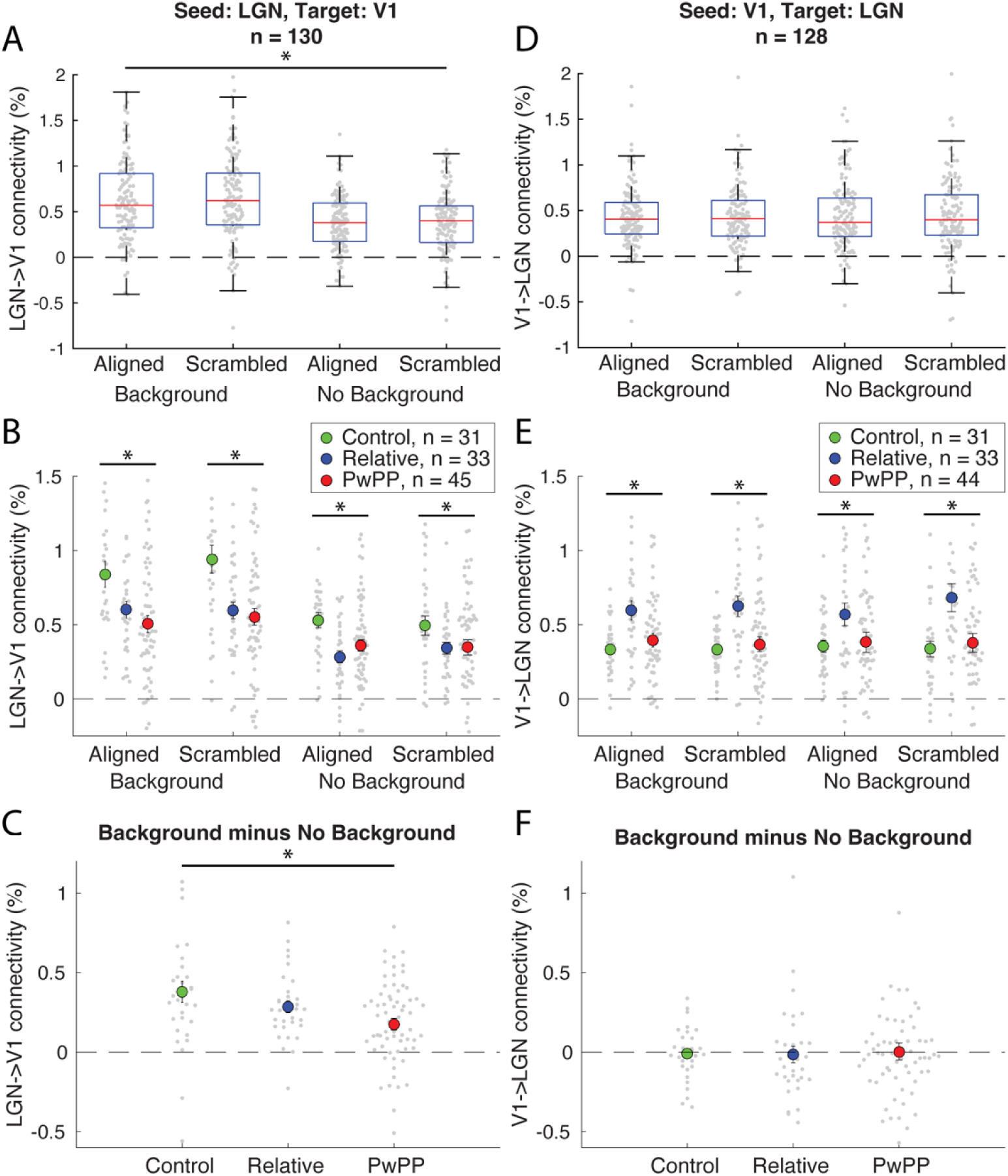
Task-based functional connectivity analysis between LGN and V1. **A-C)** Data with LGN as seed region and V1 as target region. **D-F)** Data with V1 as the seed region and LGN as the target region. In **A** & **D**, red line shows the median, boxes show interquartile range, whiskers show 1.5 x interquartile range, gray dots show all data points. In **B**, **C**, **E**, & **F**, error bars show ± 1 *SEM*. Asterisks indicate significant differences across background conditions in A, across groups in B and E, and a group by background interaction (averaging across aligned & scrambled contours) in C (*p* < 0.05).

With LGN as the seed region and V1 as the target, functional connectivity was significantly higher for contours presented within a field of background elements versus contours presented without a background (ANOVA, main effect of background; *F* _(1, 106)_ = 87.91, *p* < 0.001; *Figure 7A*). Healthy controls showed higher functional connectivity overall between LGN-V1 compared to first-degree relatives and PwPP (ANOVA, main effect of group; *F* _(2, 106)_ = 7.15, *p* = 0.001; *Figure 7B*). We also found that the difference in functional connectivity for stimuli with versus without background elements was significantly higher in healthy controls compared to individuals with psychosis, with first-degree relatives showing an intermediate response pattern (ANOVA, group by background interaction; i.e., averaging across aligned & scrambled contours; *F* _(2, 106)_ = 5.78, *p* = 0.004; *Figure 7C*). This result suggests an abnormal functional relationship between LGN and V1 among PwPP during visual contour perception, wherein LGN-V1 connectivity in the presence of background stimuli was poorly modulated in this group of participants.

With V1 as the seed region and LGN as the target, first-degree relatives showed significantly higher functional connectivity overall compared to healthy controls and individuals with psychosis across conditions (ANOVA, main effect of group; *F* _(2, 106)_ = 7.0, *p* = 0.001; *Figure 7E*). Considering lower LGN-V1 connectivity among relatives versus controls (above), higher V1-LGN connectivity may reflect a compensatory mechanism among people with a genetic liability for psychosis. No other significant main effects or interactions were seen for the V1-LGN data (ANOVA; *F*-values < 0.02, *p*-values > 0.2; *Figure 7D, E, & F*). As we did not see any significant differences in univariate fMRI responses (either V1 or LGN) between participants with schizophrenia and bipolar disorder (*Supplemental Figure 5*), we did not carry out comparisons of task based functional connectivity between these groups.

#### 3.2.5 Task-based functional connectivity between V1 and LOC

We also examined task-based functional connectivity between V1 and LOC, to explore the role of functional interactions between these two visual cortical regions during contour object perception in psychosis (*Figure 8*). With V1 as the seed region and LOC as the target, we observed no significant main effects of group, contour, or background (ANOVA, main effects; *F*-values < 6.58, *p*-values > 0.08; *Figure 8A & B*). However, we found a significant group by background interaction (*F* _(2, 104)_ = 3.28, *p* = 0.04; *Figure 8B & C*); this indicates that the presence of background stimuli led to a larger increase in V1-LOC connectivity (versus no background) for PwPP and relatives versus controls.

**Figure 8:**
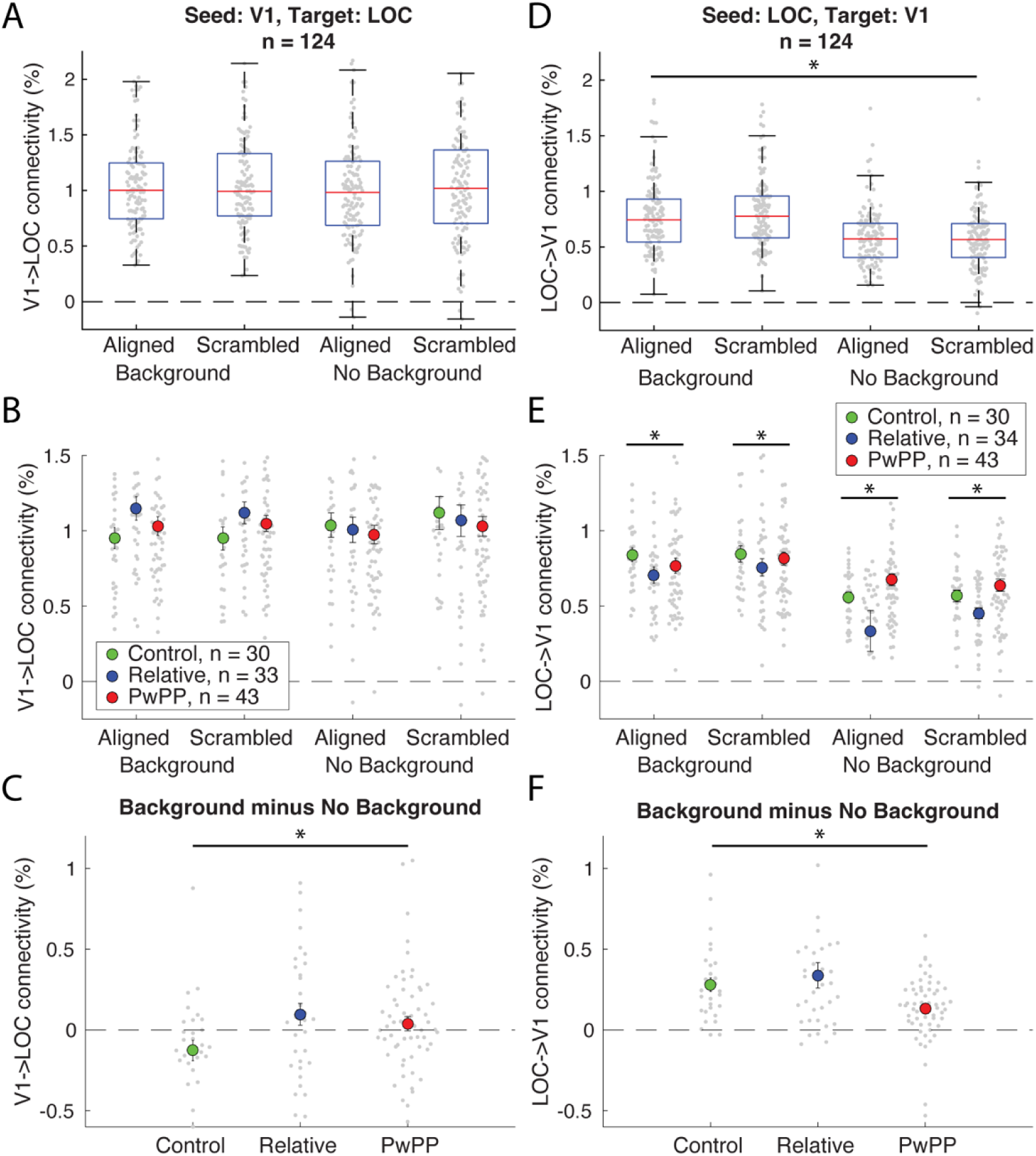
Task-based functional connectivity analysis between V1 and LOC. **A-C)** Data with LOC as seed region and V1 as target region. **D-F)** Data with V1 as the seed region and LOC as the target region. In **A** & **D**, red line shows the median, boxes show interquartile range, whiskers show 1.5 x interquartile range, gray dots show all data points. In **B**, **C**, **E**, & **F**, error bars show ± 1 *SEM*. Asterisks indicate significant differences across background conditions in D, across groups in E, and group by background interactions (averaging across aligned & scrambled contours) in C and F (*p* < 0.05).

With LOC as the seed region and V1 as the target, we saw significantly higher functional connectivity for contour stimuli presented with versus without background elements (ANOVA, main effect of background; *F* _(1, 104)_ = 71.82, *p* < 0.001; *Figure 8D*). We also saw a main effect of group, with first-degree relatives showing significantly lower functional connectivity compared to healthy controls and PwPP (ANOVA, main effect of group; *F* _(2, 104)_ = 3.2, *p* = 0.04; *Figure 8E*). Further, LOC-V1 connectivity was less strongly modulated by the presence of background stimuli among PwPP (ANOVA, group by background interaction; *F* _(2, 104)_ = 5.59, *p* = 0.005; *Figure 8E & F*). This is essentially the inverse of the pattern from the group by background interaction we observed in the above analysis with V1 as the seed and LOC as the target. Together, the results from our beta series analyses in V1-LOC reflect a failure among PwPP to modulate functional connectivity between these two regions based on the presence of background stimuli. This suggests that the functional relationship between these brain regions may play an important role in abnormal figure-ground segmentation in PwPP. As we did not see any significant differences in univariate fMRI responses (either V1 or LOC) between participants with schizophrenia and bipolar disorder (*Supplemental Figure 5*), we did not carry out comparisons of task based functional connectivity between these groups.

## 4. Discussion

We found impaired contour discrimination accuracy for aligned contours with background elements in PwPP compared to healthy controls, which was largely attributable to worse accuracy in participants with schizophrenia (*Figure 3* & *Supplemental Figure 2*). Difficulties with contour perception were also evident in lower jitter thresholds in PwPP versus biological relatives, and lower thresholds for participants with schizophrenia vs. controls and participants with bipolar disorder. Using 7T fMRI to probe the neural basis of contour perception, we observed higher responses in LOC among PwPP vs. controls (*Figure 6*), in addition to differences in task-based functional connectivity between the LGN and V1, as well as between V1 and LOC during the contour perception task (*Figure 7* & *Figure 8*).

These abnormalities were most prominent in conditions where stimuli were seen with background elements present, suggesting that contour processing deficits in PwPP emerge under conditions that require perceptual segmentation of object / figural elements from distracting background features in a visual scene. In general, fMRI responses in biological relatives were similar to those in PwPP, whereas contour discrimination task performance among relatives was normative. This suggests an influence of genetic liability for psychosis on the contributions of early visual areas to contour processing, and that compensatory mechanisms may support normative task performance among relatives. Together, our findings provide new insight into the neural basis of impaired visual integration among people with psychosis spectrum disorders by highlighting how activation within specific visual regions and their functional connectivity may contribute to these deficits.

Our behavioral data revealed poorer contour discrimination accuracy and lower (impaired) jitter thresholds among PwPP, especially those with schizophrenia. Previous studies have yielded mixed results with regard to difference in jitter thresholds during contour integration in psychosis, with some studies observing significant impairments relative to controls (Grove et al., 2018; Keane et al., 2012, 2016; Pokorny, Lano, et al., 2021; Robol et al., 2013; Schallmo et al., 2013), and others finding no such difference (Moran et al., 2022; Silverstein et al., 2012, 2015). Our results provide evidence in favor of such an impairment in jittered contour discrimination thresholds among PwPP, especially individuals with schizophrenia, and support the notion of normative contour perception among biological relatives (Pokorny, Lano, et al., 2021; Schallmo et al., 2013).

After quantifying discrimination accuracy for contours with and without background elements, we excluded participants who showed poor performance on these catch trials from further analyses, which suggests that our pattern of behavioral results cannot be explained solely by a generalized task deficit among PwPP. However, we did observe a significant correlation between jitter thresholds and BACS scores, suggesting a relationship between poorer cognitive functioning and impaired contour discrimination performance. We also saw correlations between lower thresholds and higher scores for both SGI and SPQ, suggesting contour perception may track with more subtle characteristics of psychosis (SPQ) and unusual real-world perceptual experiences (SGI). However, these last two correlations did not survive correction for multiple comparisons, and thus should be interpreted with caution.

The results from our univariate fMRI analyses replicate and extend previous work in several important directions. Previous studies have found that PwPP and healthy controls show similar levels of fMRI responses in V1 during contour discrimination tasks (Silverstein et al., 2015), and we see that this is true even when examining responses within voxels mapped to the retinotopic position of the contour stimulus. We have extended this by showing there are also no significant differences between PwPP and controls in fMRI responses from the LGN (*Figure 5*). We speculate that higher LGN responses among first degree relatives might reflect a compensatory mechanism among people with a genetic liability for psychosis during contour perception. We also observed higher fMRI responses in LOC in PwPP (*Figure 6*), consistent with prior work in the field (Silverstein et al., 2015). However, we found this difference was only present in conditions with background elements, suggesting that PwPP may exhibit impaired suppression of background distractor elements during contour integration (for a relevant EEG study of the effect of background stimuli on contour perception among PwPP, see Pokorny, Lano, et al., 2021).

The impact of background stimuli on contour perception in psychosis is further reflected in our functional connectivity analyses, wherein we saw an interesting difference in connectivity between V1 and LOC for our psychosis versus control groups. PwPP, in comparison to healthy controls, showed less modulation of V1-LOC connectivity for contours with versus without backgrounds, indicating an abnormality in the functional relationship between V1 and LOC (*Figure 8*). This difference in functional connectivity may suggest a failure to suppress background noise elements relative to contour elements (i.e., figure-ground segmentation) among PwPP. This result is broadly similar to a recent study showing differences in task-based functional connectivity across visual and frontal regions among PwPP when viewing fragmented objects (Pokorny, Espensen-Sturges, et al., 2021). However, in that study, group differences in V1-LOC connectivity were not significant.

Our functional connectivity analyses also revealed differences in the functional relationship between LGN and V1 between PwPP, their relatives, and healthy controls. Studies in animal models suggest an important role for reciprocal processing between LGN and V1 during visual contour perception (Sillito et al., 2006). In the current study, we found LGN to V1 connectivity weights were lower among PwPP versus controls, especially in the presence of background stimuli (*Figure 7*). This appears consistent with the thalamocortical dysconnectivity hypothesis in schizophrenia (Anticevic et al., 2014; Cheng et al., 2015; Damaraju et al., 2014; Dong et al., 2019; Giraldo-Chica & Woodward, 2017; Ramsay, 2019; Woodward et al., 2012), and suggests that abnormally reduced LGN-V1 connectivity might play a role in dysfunctional contour integration among PwPP. Further, V1-LGN connectivity was highest among relatives, whereas LGN-V1 connectivity was intermediate for relatives versus PwPP and controls. We speculate that this might reflect a compensatory mechanism by which reciprocal processing between LGN and V1 (Sillito et al., 2006) helps support normative contour integration among people with a genetic liability for psychotic psychopathology.

This study had some limitations, which arose from design choices implemented to enhance other facets of the study. Our focus on participants with psychotic symptoms, rather than specific diagnostic categories, necessarily reduced the number of individuals in each particular diagnostic category within our sample, which meant that we had lower statistical power to perform analyses comparing between diagnostic groups (e.g., schizophrenia vs. bipolar disorder). We took an ROI-based approach to our fMRI analyses, informed by an understanding of the role different visual areas play in contour processing. The use of visual ROIs selected *a priori* based on anatomical and functional criteria necessarily limited our examination of the role(s) played by other brain regions and networks during contour perception in PwPP. Future studies using this data set will focus on whole brain and network analyses of brain activation during contour object perception in psychosis.

This study had several strengths, the first being a relatively large sample size, and overall high data quality (for additional quantification and group comparisons of data quality, see Schallmo et al., 2023). Including first degree relatives allowed us to examine the role of genetic liability for psychosis in contour integration. Studying PwPP trans-diagnostically allowed us to assess contour perception as a function of psychotic symptoms, in addition to comparing across diagnostic categories, whose reliability and validity have been questioned (Kotov et al., 2017; Markon et al., 2011). Comparing across psychosis, relative, and control groups enabled us to assess contour processing across a spectrum of individuals who varied in their levels of clinical symptoms and visual functioning. Our use of 7T fMRI gave us a higher functional contrast-to-noise ratio and higher spatial resolution (as compared to 3T, for example), and our fMRI pre-processing pipeline included distortion compensation, gradient nonlinearity correction, motion censoring, and cluster-based ROI definitions using spatial auto-correlations, to address known artifacts. Finally, these data were acquired as part of the Psychosis Human Connectome Project, which included HCP-style acquisition of structural and resting state functional connectivity data at 3T (Demro et al., 2021), as well as other visual task fMRI and MR spectroscopy data at 7T (Schallmo et al., 2023). This large multi-modal dataset is publicly available; we hope this will facilitate additional future studies of visual contour integration in psychosis using these same data.

## Data Availability

Data for this study are available through the National Data Archive (nda.nih.gov/edit_collection.html?id=3162).

## Acknowledgements

We thank Jesslyn (Li Shen) Chong, Victoria Espensen-Sturges, and Marisa J. Sanchez for help with data collection. This work was supported by the National Institutes of Health (U01 MH108150). Salary support for MPS was provided in part by K01 MH120278. MR scanning at the University of Minnesota Center for Magnetic Resonance Research was supported by P41 EB015894 and P30 NS076408. This study used tools from the University of Minnesota Clinical and Translational Science Institute supported by UL1 TR002494.

## Supplemental Material

**Supplemental Figure 1:**
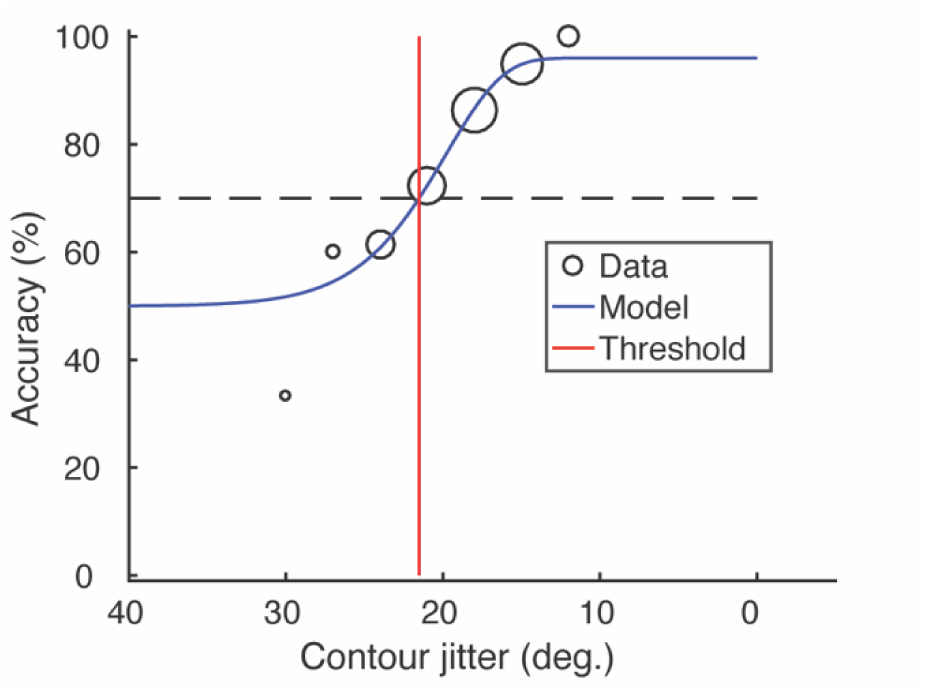
Example psychometric function. Psychophysical data (black) from a single participant were fit with a Weibull function (blue) to obtain a jitter threshold (red), which reflects 70% contrast discrimination accuracy (dashed line). Adapted from Schallmo et al. (2023).

**Supplemental Figure 2:**
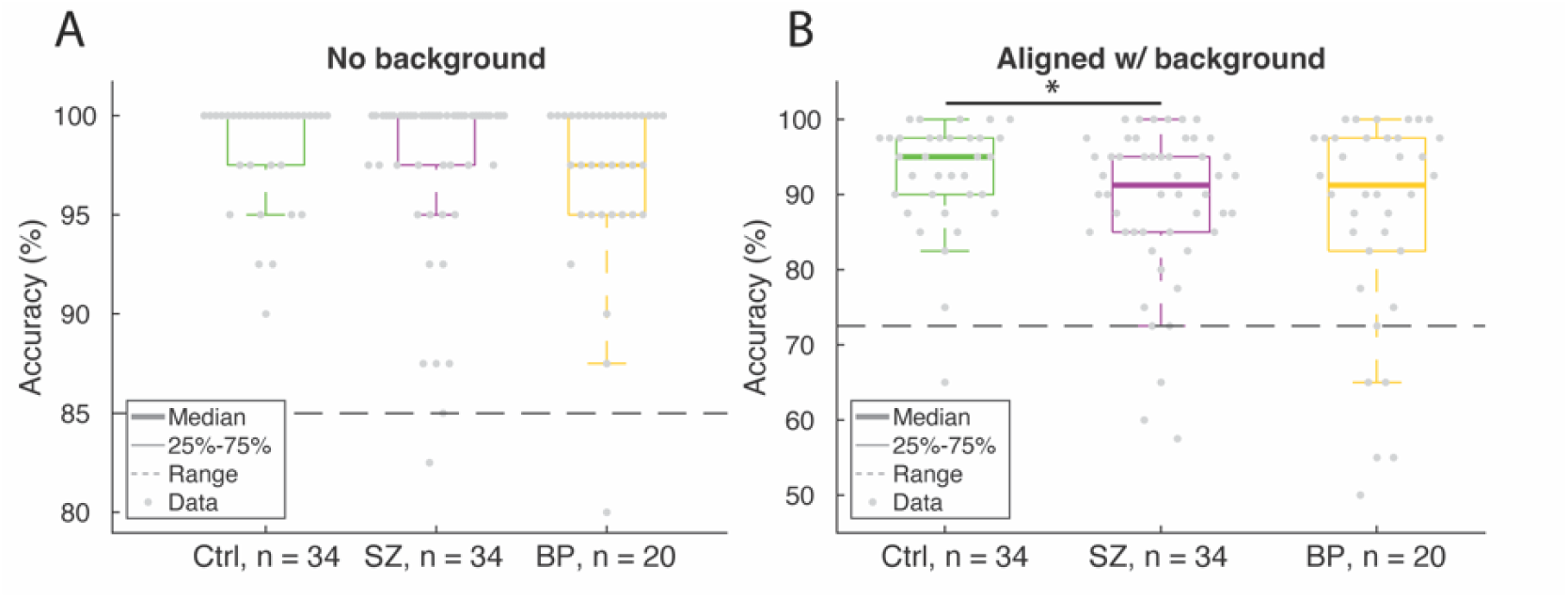
Post-hoc analyses between healthy controls, participants with schizophrenia, and participants with bipolar disorder. **A)** Contour discrimination accuracy for scrambled contour stimuli without background elements, and **B)** accuracy for aligned contour stimuli with background elements. Error bars show ± 1 *SEM*. Asterisk indicates significant difference across participant groups (*p* < 0.05).

**Supplemental Figure 3:**
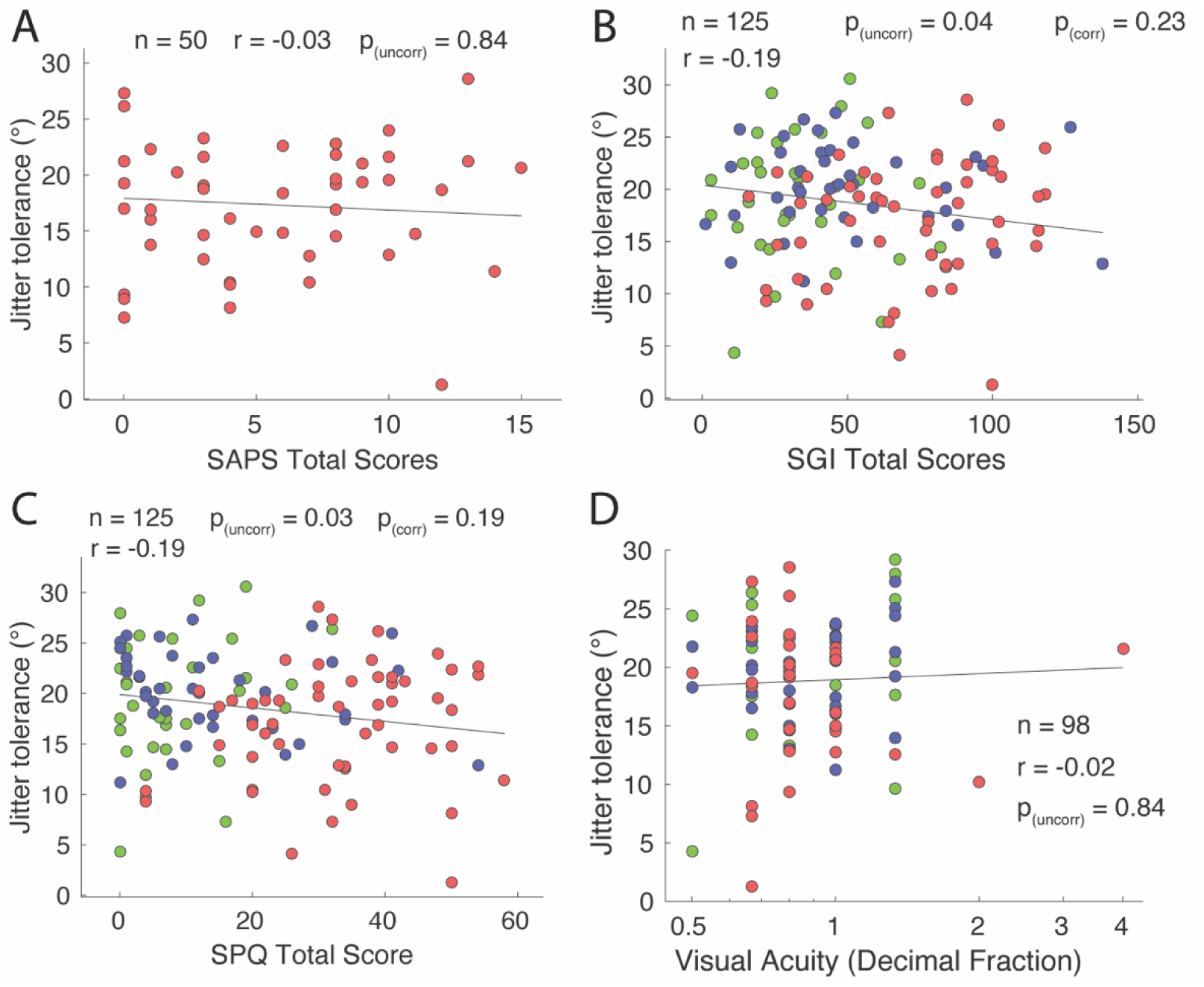
Relationship between jitter thresholds and clinical measures. **A)** SAPS Total Score. **B)** SGI Total Score. **C)** SPQ Total Score. **D)** Visual acuity measured using Snellen chart. Log scale of decimal fraction values are reported (e.g., 0.5 indicates 20/40). *r*-values are Pearson correlations (**A-C**), or Spearman ranked correlations (**D**). *p*-values are shown as uncorrected (uncorr), or Bonferroni-corrected for multiple comparisons (corr).

**Supplemental Figure 4:**
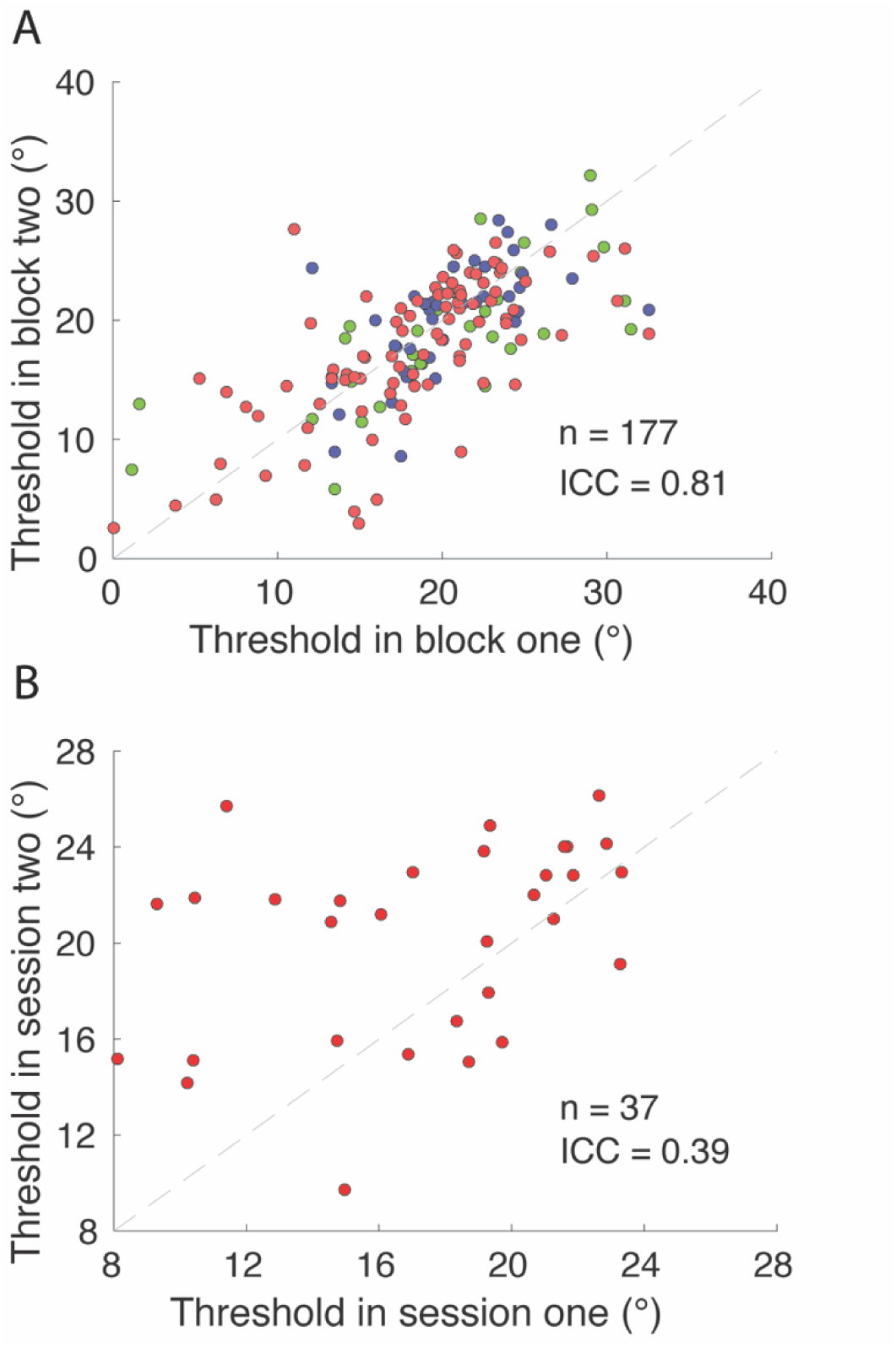
Threshold reliability measurements. **A)** Threshold estimates across the two experimental blocks of behavioral psychophysics. Points are shown for each data set, and there are more data sets than unique participants due to repeat data. **B)** Threshold estimates across the first and second sessions from the same participant. ICC = intraclass correlation coefficient (ICC (3, k); Koo, 2016).

**Supplemental Figure 5:**
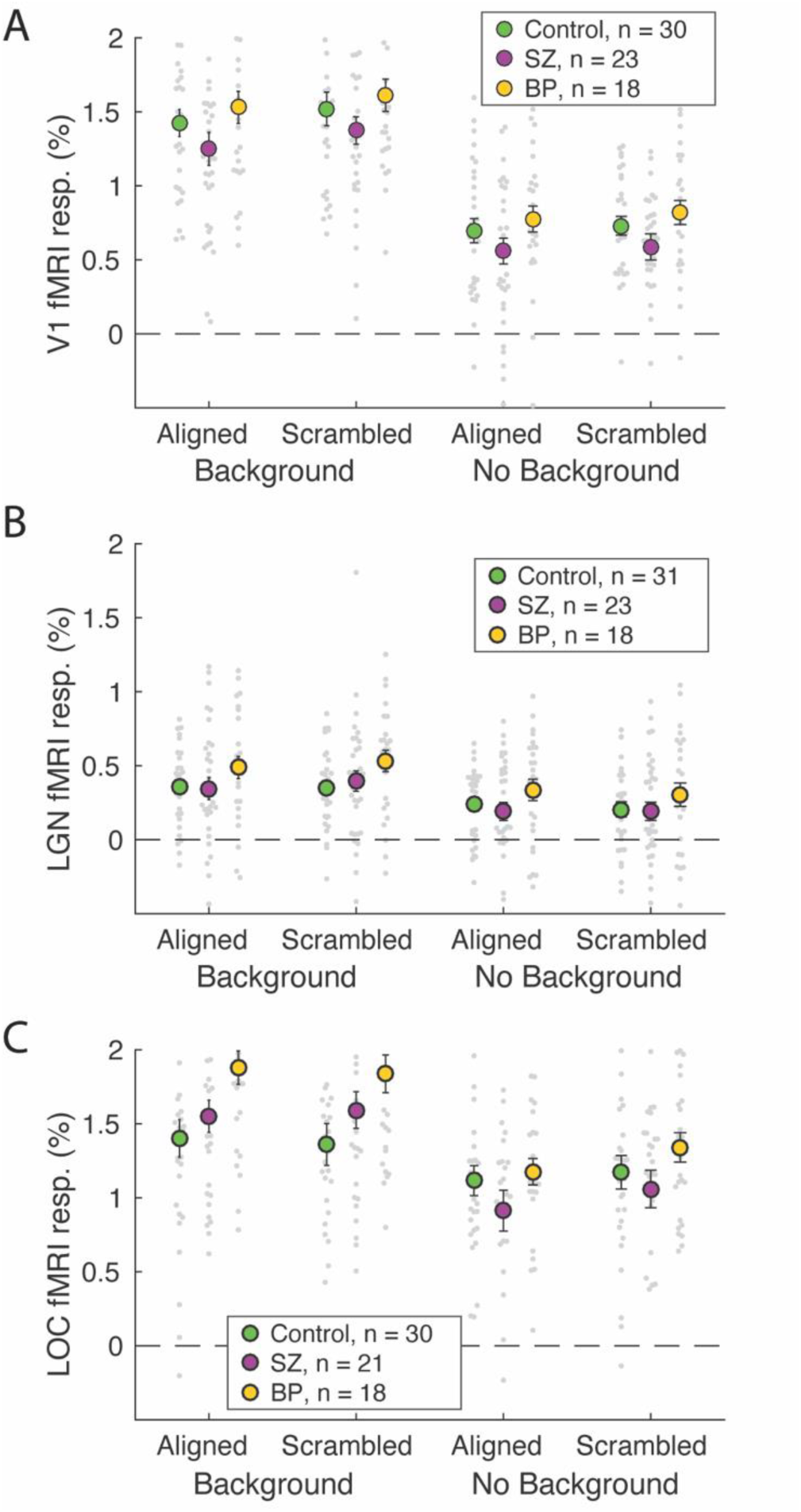
Post-hoc analyses on univariate fMRI data between healthy controls, participants with schizophrenia, and participants with bipolar disorder. **A)** V1, **B)** LGN, and **C)** LOC fMRI responses across participant groups. Small gray dots show individual subject data, larger circles show group means. Not all individual data points are shown, to better visualize the group means. Error bars show ± 1 *SEM*.

**Supplemental Table 1:** Unique participants and repeat sessions for the psychophysics portion of the study and for each ROI from fMRI across participant groups.

| Portion of study | Unique participants (Ctrl / Rel / PwPP) | Repeat sessions (PwPP only) |
| --- | --- | --- |
| Psychophysics | 34 / 44 / 62 | 37 |
| V1 ROI | 30 / 34 / 45 | 22 |
| LGN ROI | 31 / 34 / 45 | 23 |
| LOC ROI | 30 / 34 / 43 | 18 |

